# Not all roads lead to the immune system: The Genetic Basis of Multiple Sclerosis Severity Implicates Central Nervous System and Mitochondrial Involvement

**DOI:** 10.1101/2022.02.04.22270362

**Authors:** Vilija G. Jokubaitis, Omar Ibrahim, Jim Stankovich, Pavlina Kleinova, Fuencisla Matesanz, Daniel Hui, Sara Eichau, Mark Slee, Jeannette Lechner-Scott, Rodney Lea, Trevor J Kilpatrick, Tomas Kalincik, Philip L. De Jager, Ashley Beecham, Jacob L. McCauley, Bruce V. Taylor, Steve Vucic, Louise Laverick, Karolina Vodehnalova, Maria-Isabel García-Sanchéz, Antonio Alcina, Anneke van der Walt, Eva Kubala Havrdova, Guillermo Izquierdo, Nikolaos Patsopoulos, Dana Horakova, Helmut Butzkueven

## Abstract

Multiple sclerosis (MS) is a leading cause of neurological disability in adults. Heterogeneity in MS clinical presentation has posed a major challenge for identifying genetic variants associated with disease outcomes. To overcome this challenge, we used prospectively ascertained clinical outcomes data from the largest international MS Registry, MSBase. We assembled a cohort of deeply phenotyped individuals with relapse-onset MS. We used unbiased genome-wide association study and machine learning approaches to assess the genetic contribution to longitudinally defined MS severity phenotypes in 1,813 individuals. Our results did not identify any variants of moderate to large effect sizes that met genome-wide significance thresholds. However, we demonstrate that clinical outcomes in relapse-onset MS are associated with multiple genetic loci of small effect sizes. Using a machine learning approach incorporating over 62,000 variants and demographic variables available at MS disease onset, we could predict severity with an area under the receiver operator curve (AUROC) of 0.87 (95% CI 0.83 – 0.91). This approach, if externally validated, could quickly prove useful for clinical stratification at MS onset. Further, we find evidence to support central nervous system and mitochondrial involvement in determining MS severity.

---

Multiple sclerosis (MS), a complex trait disease, is a leading cause of non-traumatic neurological disability in adults. It affects approximately 2.8 million people worldwide, predominantly females.^1^ Rates of disability progression and long-term outcomes are highly heterogeneous amongst people with relapse-onset MS (RMS).^2^ At present, the ability to predict a person’s likely long-term disease outcome at onset is very limited, but highly desirable, in order to stratify individuals for initiation with the most appropriate disease-modifying therapy.

To-date, over 230 common variants have been linked to MS risk.^3^ The only replicated genetic modifier of MS phenotype is carriage of the principal risk allele, the human leukocyte antigen (HLA) DRB1*15:01. In European populations, carriage of the HLA-DRB1*15:01 allele confers younger age of onset.^4^ However, large studies have shown that this allele has no effect on MS progression after onset.^5, 6^ Further, there is strong evidence to suggest that currently known risk variants, aside from HLA-DRB1*15:01, play no major role in determining MS severity.^7-9^

A genetic influence on MS outcome is, however, plausible, in particular relating to the severity of secondary inflammation (e.g. development of slowly expanding, or chronic rim-active lesions), resilience to neuroaxonal injury, or remyelination capacity. Indeed, preliminary genome-wide association study (GWAS) evidence suggests that susceptibility and severity likely involve distinct biological processes and pathways.^10-12^

The best evidence to-date for a genetic contribution to disease outcomes comes from a small number of cross-sectional GWAS dedicated to a search for severity signals associated with the MS severity scale (MSSS) score^13^, or age at onset.^9, 10, 14-17^ However, these signals failed to reach significance at the genome-wide level, possibly due to inclusion of populations with both relapse-onset and progressive-onset clinical courses. As the genetic architecture underlying worsening in relapse-onset MS and progressive-onset MS is possibly distinct,^18^ it could be important to study these populations separately. Further, use of limited cross-sectional phenotypic MSSS data to assess disease severity limits accurate severity phenotyping due to both major ceiling effects and instability in RMS.^13^

The heterogeneity in MS severity, both between individuals, and within individuals over time, is large. Therefore, analysis of longitudinally acquired clinical trajectories over many years is likely to be more reliable for accurate severity assignation. Given that preliminary evidence suggests that genetic variation influences severity outcomes, we used both unbiased genome-wide association, and machine learning approaches to examine 1) whether prospectively ascertained, longitudinally-defined RMS phenotypes could reveal novel genetic variants associated with disease severity, 2) whether a machine learning model with multi-single nucleotide variant (SNV) inclusion has sufficient positive predictive value to potentially be used at the time of MS diagnosis to guide clinical and treatment decisions. Our secondary analyses further interrogated SNV signals derived from our primary analyses, and also aimed to replicate previously reported suggestive markers of MS severity using a targeted approach.

## Results

### Cohort characteristics

The cohort comprised of 5,851 people with relapse-onset MS from Australia, the Czech Republic, and Spain (**Figure S1**). Those who met study minimum inclusion criteria (**Figure S2**), represented 63,072 patient-years of follow-up. Of these, 1,984 (33.9%) people were genotyped, of whom 1,813 (91.4%), representing 22,884 patient-years of follow-up, passed additional filtering and genotyping quality control (QC**; Table S1**). The clinical and demographic characteristics of the cohort based on longitudinal age-related MS severity scale^19^ (l-ARMSS) scores (**Table 1**), and longitudinal MSSS (l-MSSS; **Table S2**) are shown. Per-country cohort characteristics are provided in **Table S3**. Individual phenotypes based on continuous l-ARMSS and l-MSSS, binary l-ARMSS and l-MSSS, Age at Onset (AAO), and MS susceptibility weighted genetic risk scores (wGRS) are available in **Table S4**. The correlation between l-ARMSS and l-MSSS was strong (r=0.90, p<0.0001, **Table S4**). l-ARMSS and l-MSSS scores in individual disease trajectories are shown in **Figure 1**.

**Table 1:**
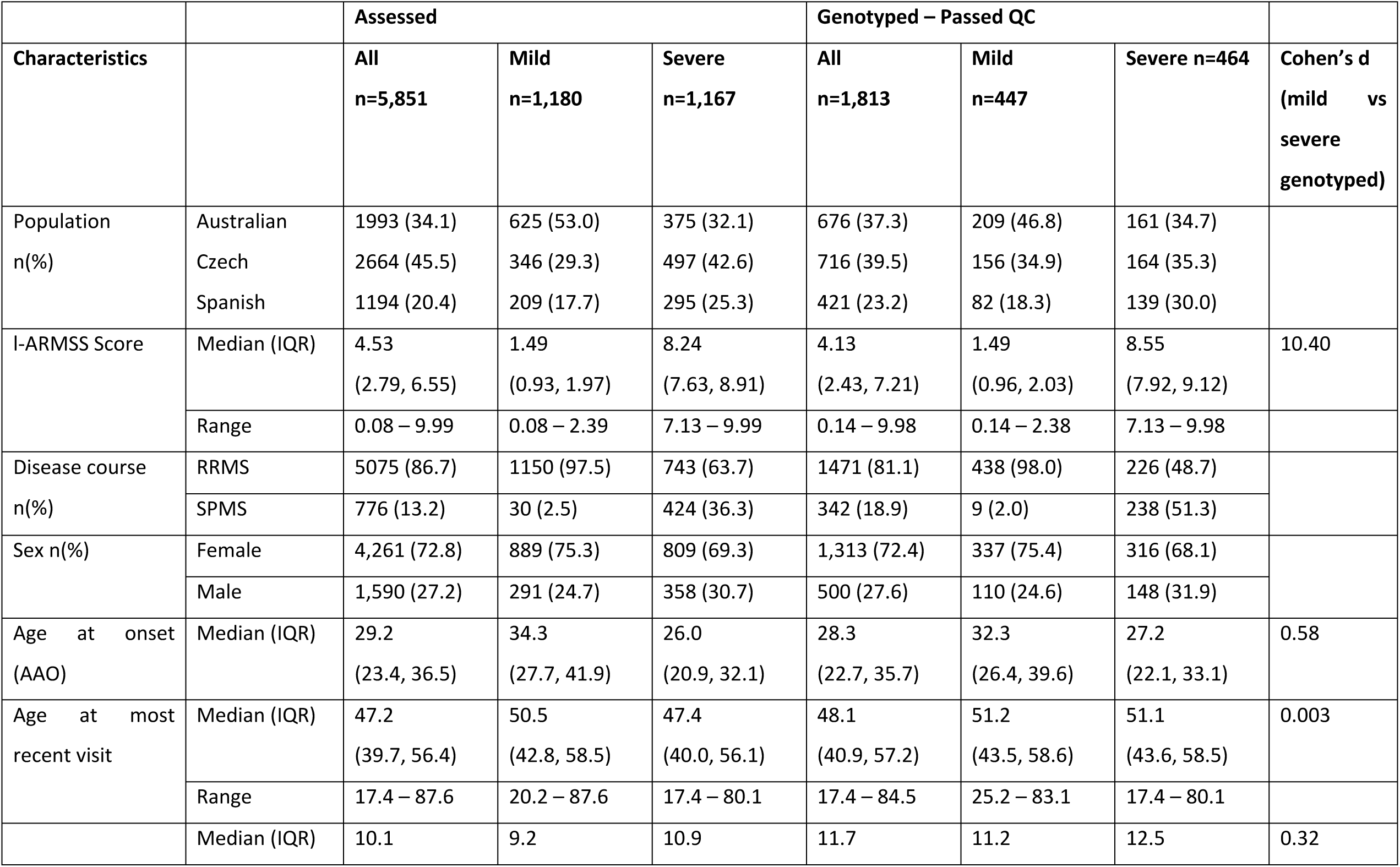
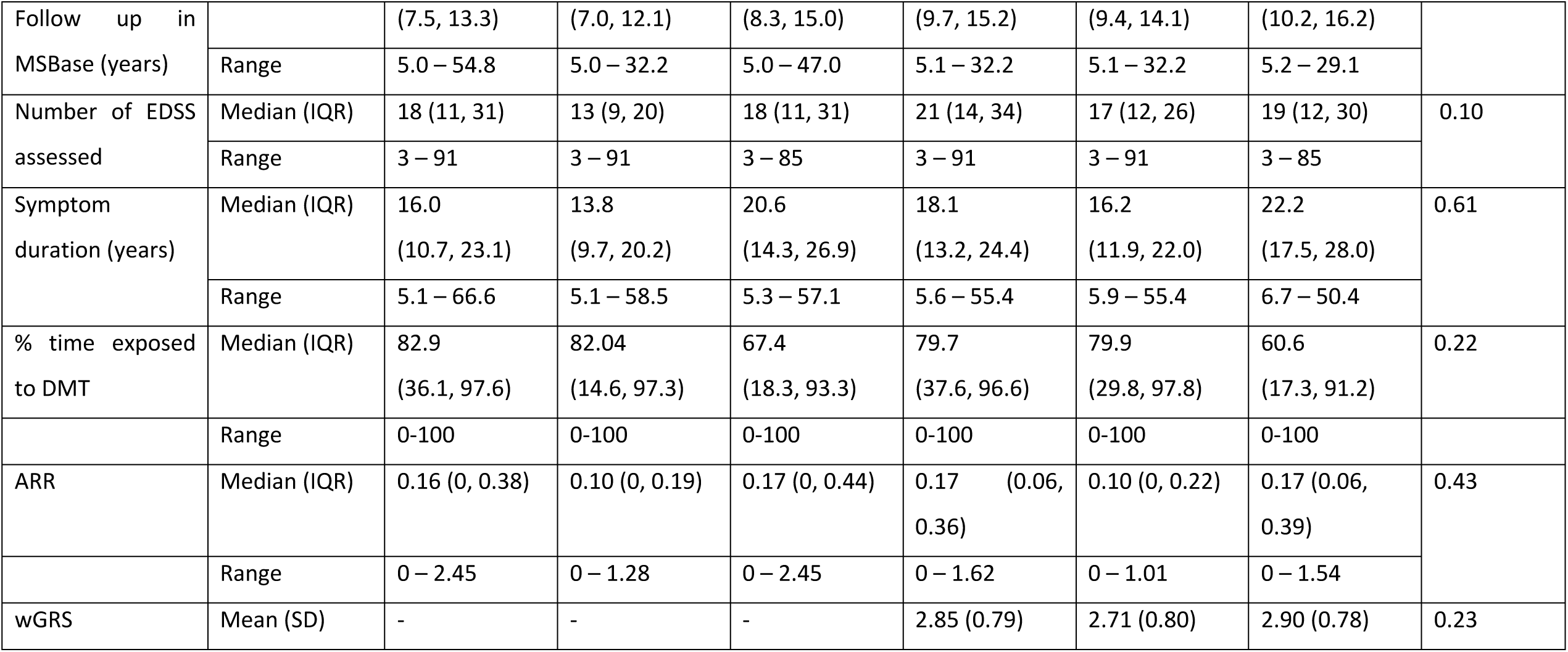
Cohort Characteristics by longitudinal ARMSS (l-ARMSS) categorisation.

**Figure 1:**
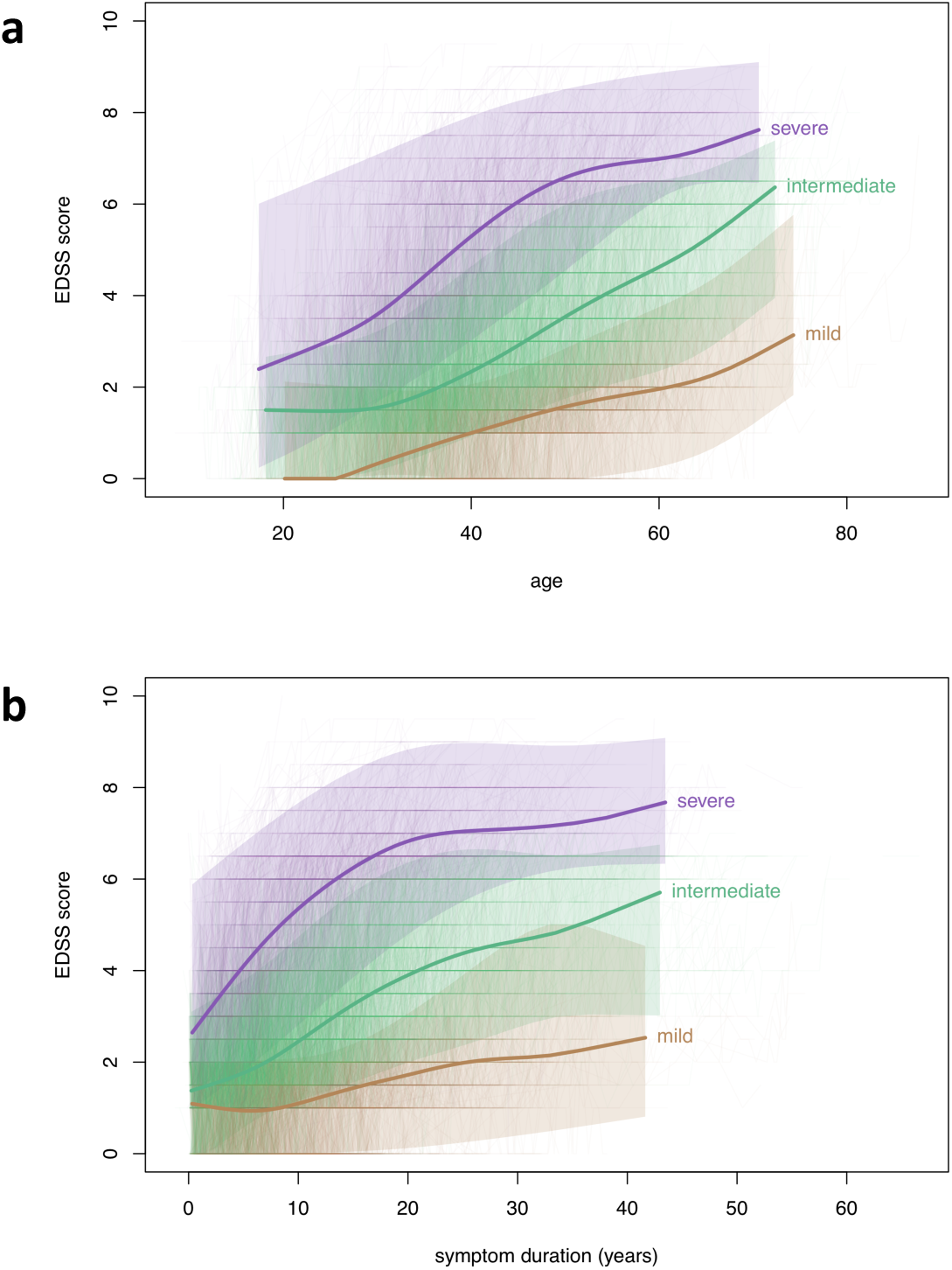
EDSS-age trajectories for included participants classified into mild (brown; n=1,180), intermediate (green; n=3,559) or severe (purple; n=1,112) groups based on median longitudinal ARMSS scores. **1b:** EDSS-symptom duration trajectories for included participants classified into mild (brown; n=1,232), intermediate (green; n=3,512), or severe (purple; n=1,107) groups based on median longitudinal MSSS scores.

### Primary analyses

#### Genome-wide association search for SNVs associated with longitudinal severity measures

We first performed a *genome-wide association* analysis to identify novel variants associated with l-ARMSS continuous (**Table S5**) or binary (**Table S6**) phenotypes. Cohort characteristics described in **Table 1** demonstrated that those in the severe l-ARMSS cohort had longer follow up (12.5 years v 11.2 years, Cohen’s d = 0.32), longer symptom duration (22.2 years v 16.2 years, Cohen’s d=0.61), a younger age at onset (27.2 years vs 32.3 years, Cohen’s d =0.58), a higher annualised relapse rate (0.14 v 0.10, Cohen’s d=0.43), a lower cumulative proportion of time exposed to disease-modifying therapy (60.6% vs 79.9%, Cohen’s d=0.22), and a higher wGRS (2.90 vs 2.71, Cohen’s d=0.23) relative to the mild cohort. Therefore, all regression analyses of l-ARMSS phenotypes were *a priori* adjusted for the first 5 principal components (PCs), the number of HLA-DRB1*15:01 alleles carried, percentage of time exposed to disease-modifying therapy and imbalanced variables as above. Fixed-effects meta-analysis results from six groups (two from each country) did not identify any single nucleotide variants (SNVs) that surpassed genome-wide significance (p<5×10^−8^) for any of the above phenotypic outcomes (**Table S5; Figure S3**). Similarly, adjusted I-MSSS analyses (**Table S2**) did not identify any significant associations, related to continuous (**Table S7**; **Figure S4**), or binary (**Table S8**) phenotypes. Assessment of the genomic locations of SNVs with p<1×10^−5^ for the l-ARMSS phenotype demonstrated that 55.1% (p=6.63×10^−3^ for enrichment) of the signals were in intergenic regions and 34.7% were intronic (**Figure S5a**). This was numerically different to the I-MSSS endpoint analysis, where 47.9% of SNVs were intronic (p=5.41×10^−3^ for enrichment) and 36.8% were intergenic (p=0.0193; **Figure S5b**).

A summary of the top variants associated with the continuous l-ARMSS and l-MSSS analyses are shown in **Table 2**. The top signal in the continuous l-ARMSS analysis was rs7289446 (β=-0.4882, p=2.73×10^−7^), intronic to *SEZ6L*, a gene associated with dendritic spine density and arborization.^20^ The top signal in the continuous l-MSSS analysis also implicated a variant intronic to *SEZ6L*, rs1207401 (β=-0.4841, p=2.88×10^−7^). These two SEZ6L associated SNVs are in perfect linkage disequilibrium (R^2^=1, D’=1; **Table S9; Figure S6**).

**Table 2:** Results of fixed effects meta-analyses: top SNVs for the 10 nearest genes for l-ARMSS and l-MSSS phenotypes & top SNVs for the 5 nearest genes in sex-stratified analyses.

| rsID | Chr | ranked order<br>SNV | ranked order<br>nearest gene | Nearest gene | Minor Allele | MAF | adj p# ^ | $\beta$ |
| --- | --- | --- | --- | --- | --- | --- | --- | --- |
| **l-ARMSS<sup>#</sup>, n=1813** |  |  |  |  |  |  |  |  |
| rs7289446 | 22 | 1 | 1 | SEZ6L | G | 0.27 | 2.73E-07 | -0.488 |
| rs1207401 | 22 | 2 | 1 | SEZ6L | A | 0.27 | 2.90E-07 | -0.490 |
| rs7758683 | 6 | 6 | 2 | EPHA7 | T | 0.23 | 1.88E-06 | -0.477 |
| rs56089601 | 4 | 7 | 3 | STK32B | C | 0.10 | 2.69E-06 | 0.655 |
| rs12953974 | 18 | 10 | 4 | CTIF | A | 0.12 | 3.64E-06 | -0.607 |
| rs56194930 | 9 | 11 | 5 | ACO1 | G | 0.11 | 3.69E-06 | 0.648 |
| rs73091975 | 7 | 14 | 6 | CCDC129 | G | 0.09 | 3.84E-06 | -0.680 |
| rs2274333 | 1 | 16 | 7 | CA6 | G | 0.29 | 4.60E-06 | -0.429 |
| rs295254 | 9 | 20 | 8 | RCL1 | G | 0.38 | 5.64E-06 | 0.388 |
| rs11057374 | 12 | 21 | 9 | DNAH10 | G | 0.35 | 5.79E-06 | -0.390 |
| rs111399831 | 13 | 29 | 10 | SUCLA2 | A | 0.21 | 7.34E-06 | -0.468 |
| **l-MSSS<sup>^</sup>, n=1813** |  |  |  |  |  |  |  |  |
| rs1207401 | 22 | 1 | 1 | SEZ6L | A | 0.27 | 2.88E-07 | -0.484 |
| rs7289446 | 22 | 4 | 1 | SEZ6L | G | 0.27 | 3.35E-07 | -0.479 |
| rs9643199 | 8 | 6 | 2 | MTSS1 | A | 0.26 | 1.90E-06 | 0.465 |
| rs2725556 | 15 | 22 | 3 | UNC13C | A | 0.08 | 2.39E-06 | 0.709 |
| rs61578937 | 8 | 23 | 4 | NCOA2 | G | 0.13 | 2.86E-06 | -0.562 |
| rs7758683 | 6 | 27 | 5 | EPHA7 | T | 0.23 | 3.61E-06 | -0.456 |
| rs56089601 | 4 | 32 | 6 | STK32B | C | 0.10 | 4.65E-06 | 0.631 |
| rs4495680 | 1 | 33 | 7 | RGS13 | G | 0.41 | 5.65E-06 | -0.390 |
| rs56363129 | 10 | 42 | 8 | SEPHS1 | G | 0.15 | 6.56E-06 | 0.516 |
| rs111399831 | 13 | 47 | 9 | SUCLA2 | A | 0.15 | 7.07E-06 | -0.534 |
| **I-ARMSS Females only, n=1313** |  |  |  |  |  |  |  |  |
| rs1219732 | 10 | 1 | 1 | CPXM2 | T | 0.35 | 6.48E-08 | 0.569 |
| rs10967273 | 9 | 3 | 2 | LOC100506422 | T | 0.13 | 1.16E-07 | 0.799 |
| rs4572384 | 16 | 15 | 3 | CRISPLD2 | A | 0.40 | 2.77E-06 | 0.487 |
| rs2776741 | 1 | 16 | 4 | RCAN3AS | A | 0.31 | 2.98E-06 | -0.516 |
| rs61873874 | 10 | 18 | 5 | MKI67 | A | 0.05 | 3.39E-06 | 1.007 |
| **I-ARMSS Males only, n=500** |  |  |  |  |  |  |  |  |
| rs7315384 | 12 | 1 | 1 | STAB2 | C | 0.24 | 1.29E-07 | 1.041 |
| rs7070182 | 10 | 2 | 2 | TCF7L2 | C | 0.18 | 3.65E-07 | -1.112 |
| rs11845242 | 14 | 4 | 3 | LINC00520 | G | 0.41 | 5.63E-07 | -0.800 |
| rs3885012 | 12 | 5 | 4 | PHLDA1 | G | 0.06 | 1.19E-06 | -1.499 |
| rs11665069 | 18 | 7 | 5 | FHOD3 | C | 0.41 | 1.34E-06 | 0.755 |
| **I-MSSS Females only, n=1313** |  |  |  |  |  |  |  |  |
| rs10967273 | 9 | 1 | 1 | LOC100506422 | T | 0.13 | **3.52E-08** | **0.830** |
| rs9643199 | 8 | 2 | 2 | MTSS1 | A | 0.26 | 6.54E-08 | 0.631 |
| rs17169210 | 5 | 4 | 3 | SLC25A48 | C | 0.23 | 1.25E-07 | -0.621 |
| rs1219732 | 10 | 7 | 4 | CPXM2 | T | 0.35 | 1.70E-07 | 0.550 |
| rs61873874 | 10 | 50 | 5 | MKI67 | A | 0.05 | 1.45E-06 | 1.051 |
| **I-MSSS Males only, n=500** |  |  |  |  |  |  |  |  |
| rs698805 | 2 | 1 | 1 | CAMKMT | G | 0.07 | **4.35E-08** | **-1.540** |
| rs7315384 | 12 | 2 | 2 | STAB2 | C | 0.24 | 8.00E-08 | 1.020 |
| rs3885012 | 12 | 14 | 4% | PHLDA1 | G | 0.06 | 3.69E-07 | -1.312 |
| rs7070182 | 10 | 35 | 5 | TCF7L2 | C | 0.18 | 3.86E-07 | -1.054 |
| rs28442172 | 18 | 51 | 6 | FHOD3 | G | 0.13 | 2.13E-06 | -1.105 |

Fixed-effects meta-analyses (n=6) adjusted for the first 5 principal components (PCs)

#### Heritability analyses

To estimate the extent to which the variability of l-ARMSS or l-MSSS-defined severity could be explained by genetic architecture, we calculated narrow-sense heritability estimates (*h*^*2*^*g*) for our cohort (n=1,813). The overall heritability estimate for the l-ARMSS phenotype was *h*^*2*^*g* 0.19 (SE 0.15, p=0.02) using the GREML by GCTA method. Similar estimates were achieved using alternate heritability estimate tools (**Table S10**). The overall l-MSSS *h*^*2*^*g* heritability estimate was slightly greater than for l-ARMSS (*h*^*2*^*g* 0.29; SE 0.14, p=0.001). However, alternate heritability estimates for l-MSSS proved highly inconsistent (**Table S10**).

#### Machine Learning

Given that, as expected, our unbiased GWAS approach did not identify any SNVs that surpassed GWAS significance thresholds, we implemented a non-linear, xgboost^21^ machine learning (ML) algorithm to determine whether a non-linear ML model could find genetic associations with severity as compared to traditional GWAS approaches. We input all SNVs with an l-ARMSS GWAS p<0.01, accounting for 62,351 variants. However, no single variant was given a weight of greater than 0.005, confirming that no genetic variant contributed appreciably to MS severity.

#### Prediction of clinical course using machine learning

Recognising that our ML model did not further illuminate the underlying genetic architecture of MS severity, we further sought to determine whether it could be used to predict severity, based on l-ARMSS outcome extremes (n=447 mild, n=464 severe). Our ML algorithm trained on 70% (n=638 l-ARMSS) of the cohort, then tested the remaining 30% (n=273 l-ARMSS). Our ML classification algorithm had high predictive accuracy, with an area under the receiver operating characteristic curve (AUROC) 0.85 (95% CI 0.80 – 0.89). The addition of AAO together with MS susceptibility wGRS (**Table S4**) further boosted the ML AUROC to 0.87 (95% CI 0.83 – 0.91; **Figure 2**). Our classification algorithm had 86% sensitivity, and 88% specificity, with a positive predictive value (PPV) of 89% and negative predictive value (NPV) of 85%. Severity classification based on l-MSSS phenotype was weaker, with an AUROC of 0.85 (95% CI 0.80-0.89), 98% sensitivity, but only 68% specificity; with a PPV of 76% and NPV of 97% (**Figure 2**).

**Figure 2:**
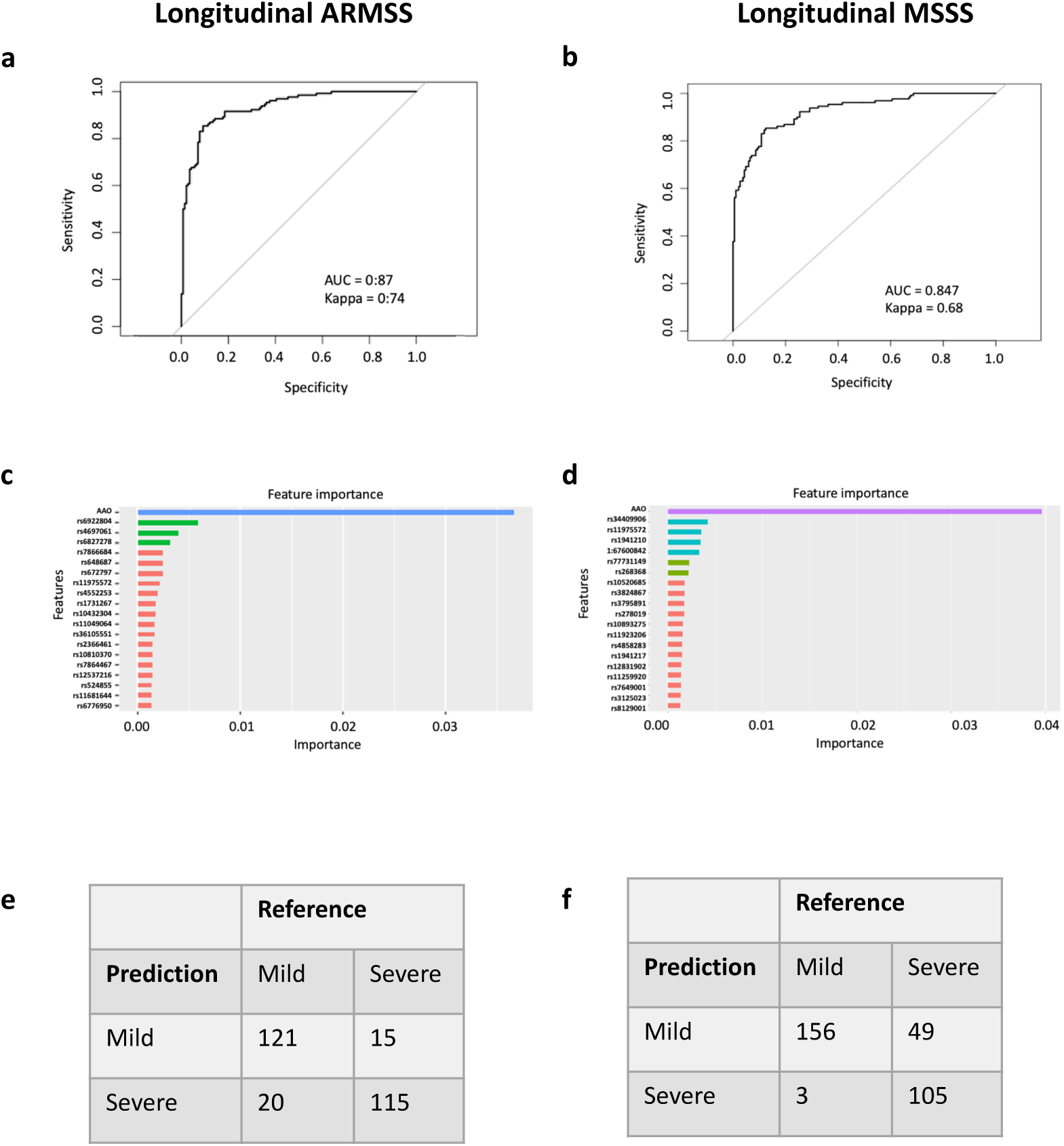
Machine learning algorithm classification of mild and severe cases. **a:** l-ARMSS +wGRS + AAO ML (n=447 mild, n=464 severe); AUROC 0.87 (95% CI 0.83-0.91) **b:** l-MSSS +wGRS + AAO ML (n=585 mild, n=466 severe); AUROC 0.85 (95% CI 0.80-0.89) **c:** Feature importance for l-ARMSS +wGRS + AAO ML model **d:** Feature importance for l-MSSS +wGRS + AAO ML model **e:** l-ARMSS +wGRS + AAO ML confusion matrix (30% cohort) **f:** l-ARMSS +wGRS + AAO ML confusion matrix (30% cohort).

Restricting the ML algorithm to just those SNVs with p<1×10^−5^ in the l-ARMSS GWAS (n=336) decreased predictive accuracy to AUROC =0.79 (95% CI 0.74 – 0.84) confirming the polygenic nature of the genetic architecture underlying MS severity.

### Secondary analyses

#### Sex-stratified Genome-wide association search for SNVs associated with longitudinal severity measures

Given our primary analyses did not identify signals of genome-wide significance, we performed sex-stratified analyses to determine whether any variant effects were potentially sex-associated. **Table 2** summarises the SNVs nearest the top 5 gene regions for each sex. The top hit in the female l-ARMSS analysis, rs1219732 intronic to *CPXM2* (β_female_ =0.5693, p=6.48×10^−08^), approached genome-wide significance (**Table S11**). This variant also approached significance in association with l-MSSS (β_female_ =0.5447, p=1.89×10^−07^, **Table S12**). We also found rs10967273, an intergenic variant, was associated with l-MSSS-defined severity in females (β_female_ =0.8289, p=3.52×10^−08^, **Table S12**; **Figure S7**). However, this variant did not surpass significance thresholds (β_female_ =0.7994, 1.17×10^−07^) in the l-ARMSS analysis.

In males, the top hit in the l-ARMSS analysis was rs7315384, intronic to *STAB2* (β_male_= 1.04, p=1.29×10^−07^), followed by rs7070182, intronic to *TCF7L2* (β_male_ =-1.11, p=3.65×10^−07^; **Table S13**). The l-MSSS analysis in males identified rs698805 intronic to *CAMKMT* (β_male_ =-1.5395, p=4.35×10^−08^) as associated with severity (**Table S14**; **Figure S8**). This variant did not surpass GWAS thresholds in the l-ARMSS analysis (β_male_ =-1.4199, p=5.64×10^−06^, **Table S13**). The variants identified in our sex stratified analyses were not associated with severity in the opposite sex (**Table S15**). We did not identify any novel genetic associations with age at onset (**Table S16; Figure S9**).

#### Pathway analyses

To identify potential biological processes overrepresented in our analyses, we analysed the top suggestive SNVs (p< 1×10^−5^) using gene-set enrichment analyses. We used FUMA^22^ to assign SNVs to genes and tissues, and genes to functions. Tissue enrichment implemented in FUMA revealed an over representation of cerebellar cortex-expressed genes for both l-ARMSS (cerebellar hemisphere p=0.071; cerebellum p=0.077; **Figure S10a**) and l-MSSS (cerebellar hemisphere p=0.017; cerebellum p=0.023); **Figure S10b**). In contrast, whole blood-associated genes were not enriched in our analyses with either l-ARMSS (p=0.75) or l-MSSS (p=0.82) outcomes. Gene set enrichment analyses of the l-ARMSS phenotype implicated endothelial cell development (β=0.43; p=8.25×10^−05^), pseudopodium assembly (β=0.84; p=2.37×10^−04^), response to progesterone (β=0.35; p=2.74×10^−04^) and NMDA receptor activity (β=1.10; p=2.79×10^−04^). The l-MSSS phenotype was additionally enriched for Wnt signalling pathways (β=0.24; p=2.02×10^−04^; **Table S17)**. We also examined gene set enrichment using Panther. Here we corroborated an overrepresentation of heteromeric G-protein signalling pathways associated with l-ARMSS (p = 4.98×10^−05^, FDR = 8.23x-10^−03^) and l-MSSS (p = 1.00×10^−04^, FDR = 1.67×10^−02^) phenotypes. The AAO phenotype was associated with endothelin (p = 2.51×10^−04^, FDR = 4.18×10^−02^), and cadherin signalling pathways (p = 2.90×10^−04^, FDR = 2.42×10^−02^).

#### Survival Analyses

Given our primary GWAS analyses did not reveal SNVs that surpassed the genome-wide level of statistical significance, we assessed whether 30 of the top signals (**Table 2**) might play a role in severity modulation using an alternative definition of severity, making use of our longitudinal dataset. Here we assessed the time to reach the hard disability milestones of irreversible expanded disability status scale (EDSS) score 3 (irEDSS3) and irreversible EDSS 6 (irEDSS6) in both univariable and adjusted analyses (**Table S18**). We identified four SNVs that were associated with time to reach irreversible EDSS 3 and 6 in both unadjusted and adjusted analyses including: rs7289446 (intronic to *SEZ6L*), rs295254 (intronic to *RCL1*), rs111399831 (nearest to *SUCLA2*), rs61578937 (nearest to *NCOA2*). These SNVs were then combined in multivariable analyses to determine whether they could independently predict time to disability milestones (**Table S19**). Three SNVs remained independently predictive of both time to irreversible EDSS 3 and 6 (**Figure 3**), including rs7289446 (*SEZ6L*: irEDSS3 adjusted HR (aHR) 0.77, p=0.008, **Figure 3a**; irEDSS6 aHR 0.72, p=4.85×10^−4^, **Figure 3b**), rs295254 (*RCL1*: irEDSS3 aHR 1.33, p=9.10×10^−4,^ **Figure 3c**; irEDSS6 aHR 1.32, p=7.37×10^−4^, **Figure 3d**) and rs111399831 (irEDSS3 aHR 0.77, p=0.036, **Figure 3e**; irEDSS6 aHR 0.62, p=2.82×10^−4^, **Figure 3f**).

**Figure 3:**
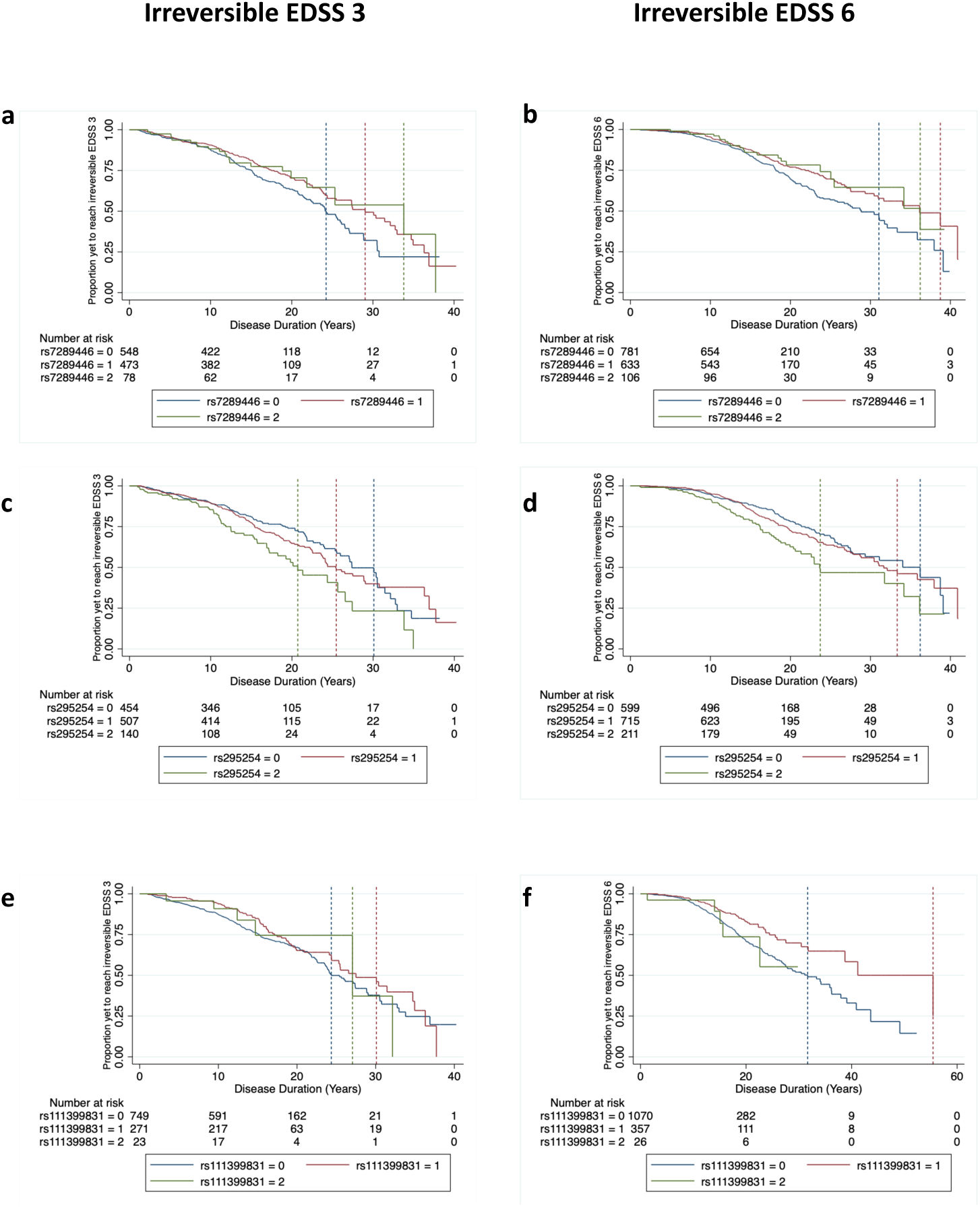
Kaplan-Meier survival curves showing time to irreversible EDSS milestones based on the presence (1,2) or absence (0) of the minor allele at each locus **a:** rs7289446 intronic to SEZ6L time to irreversible EDSS 3 (aHR 0.77, p=0.008) **b:** rs7289446 intronic to SEZ6L time to irreversible EDSS 6 (aHR 0.72, p=4.85×10^−4^) **c:** rs295254 intronic to RCL1 time to irreversible EDSS 3 (aHR 1.33, p=9.10×10^−4^) **d:** rs295254 intronic to RCL1 time to irreversible EDSS 6 (aHR 1.32, p=7.37×10^−4^) **e:** rs11399831 nearest to SUCLA2 time to irreversible EDSS 3 (aHR 0.77, p=0.036) **f:** rs11399831 nearest to SUCLA2 time to irreversible EDSS 6 (aHR 0.74, p=2.82×10^−4^).

In the sex-stratified analyses, we identified rs9643199 (intronic to *MTSS1*) and rs2776741 (nearest to *RCAN3AS*) as consistently associated with time to irreversible EDSS 3 and 6 in females (**Figure 4**), but not males (**Table S18, Figure S9**). The independent hazards of time to reach irreversible EDSS 3 and 6 for these variants were: rs9643199 (*MTSS1*: irEDSS3 aHR 1.35, p=0.006, **Figure 4a**; irEDSS6 aHR 1.46, p=7.09×10^−4^, **Figure 4b**) and rs2776741 (irEDSS3 aHR 0.77, p=0.010, **Figure 4c**; irEDSS6 aHR 0.74, p=0.005, **Figure 4d**; **Table S19**). rs7070182 intronic to *TCF7L2* (**Figure 4**) was the only variant consistently associated with time to irreversible EDSS 3 (aHR 0.59, p=0.013, **Figure 4e**) and 6 (aHR 0.56, p=0.005, **Figure 4f**) in males, with no effect in females (**Table S18**; **Figure S11**).

**Figure 4:**
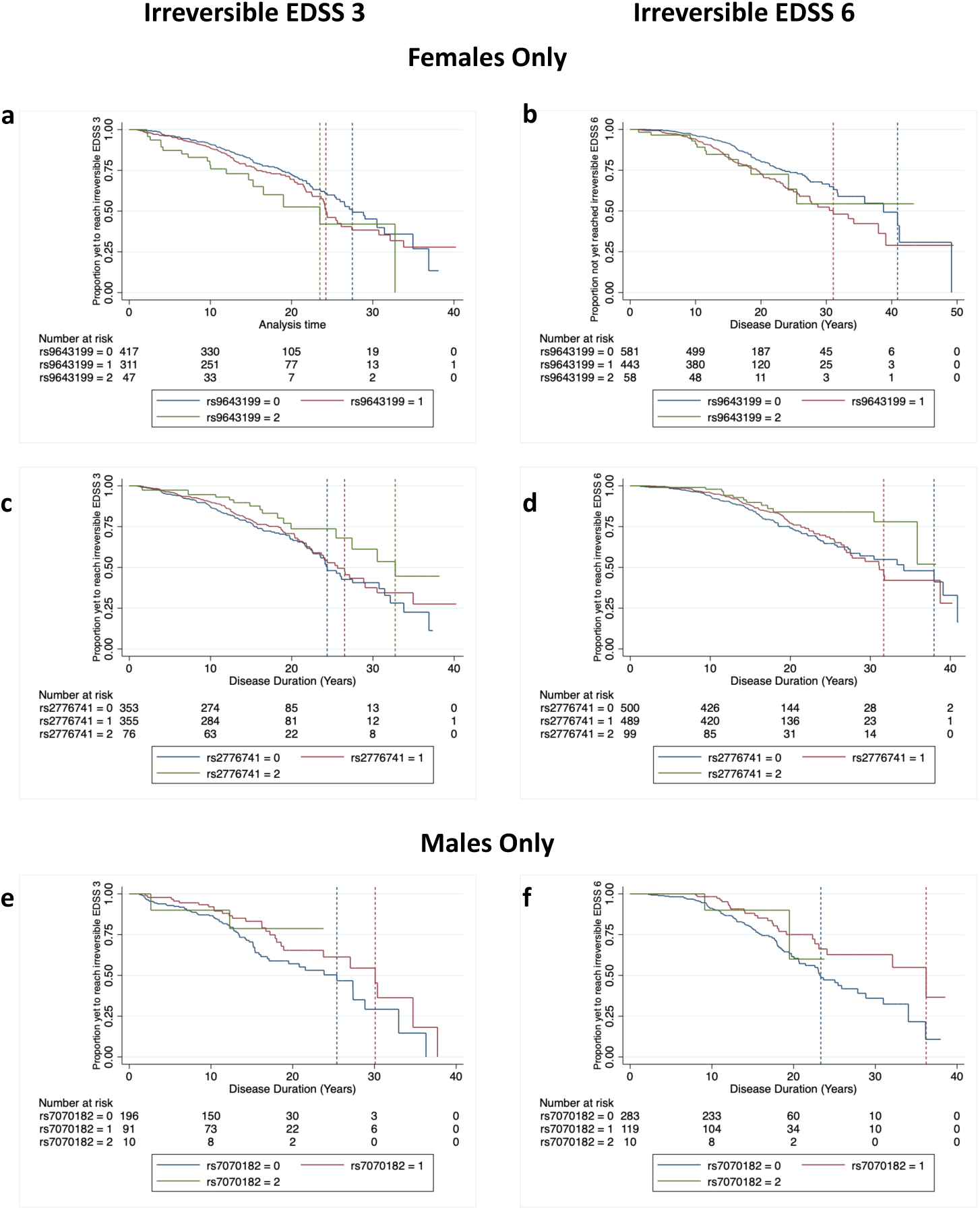
Kaplan-Meier survival curves showing time to irreversible EDSS milestones based on the presence (1,2) or absence (0) of the minor allele at each locus **a:** rs9643199 intronic to MTSS1 time to irreversible EDSS 3 in females (aHR 1.35, p=0.006) **b:** rs9643199 intronic to MTSS1 time to irreversible EDSS 6 in females (aHR 1.46, p=7.09×10^−4^) **c:** rs2776741 nearest to RCAN3AS time to irreversible EDSS 3 in females (aHR 0.77, p=0.010) **d:** rs2776741 nearest to RCAN3AS time to irreversible EDSS 6 in females (aHR 0.74, p=0.005) **e:** rs7070182 intronic to TCF7L2 time to irreversible EDSS 3 in males (aHR 0.59, p=0.013) **f:** rs7070182 intronic to TCF7L2 time to irreversible EDSS 6 in males (aHR 0.56, p=0.005).

#### MS susceptibility allele association with severity phenotypes

We sought to determine whether there was an association between known MS susceptibility variants (wGRS), and our severity phenotypes of interest. We found weak positive correlations between the MS susceptibility wGRS (**Table S20**) and l-ARMSS (r=0.07, p=0.003, **Figure S12a**); l-MSSS (r=0.03, p=0.19 **Figure S12b**); and a weak negative correlation with AAO (r=-0.08, p=0.0005) (**Figure S12c**). We did not find an association between l-AMRSS or l-MSSS and the known non-HLA autosomal risk variants^3^ that were directly genotyped (198/200), p>1×10^−3^ (**Table S20**).

The distribution, per phenotype, of HLA MS susceptibility tagging SNVs including HLA-DRB1*15:01, HLA-DRB1*03:01, HLA-DRB1*130:01, HLA-DRB1*08:01, HLA-DQB1*03:02, and protective alleles: HLA-A*02:01, HLA-DQA1*01:01, HLA-B*44:02 and HLA-B*55:01 is described in **Table S21**. We confirmed that HLA-DRB1*15:01 homozygosity was associated with an earlier AAO (rs3135388, 29.2 years v 30.4 years, p=0.005). However, homozygosity at HLA-DRB1*15:01 was not associated with disease severity as per l-ARMSS, nor l-MSSS, nor was any other SNV-genotyped HLA allele (**Table S21**).

#### Validation assessment of previously published putative severity SNVs

In addition to the main European DRB1*15:01 tagging SNV, rs3135388, we tested 116 putative non-HLA SNV associations with cross-sectional MSSS measures, disease severity, and AAO. We were able to replicate the association between rs868824, intronic to *IMMP2L* on chromosome 7, with AAO (β = -1.0935 years; p=4.31×10^−4^), however, no other putative severity variant met or surpassed the Bonferroni-corrected replication threshold (p=4.31×10^−4^, **Table S22**).

Finally, we tested the association between a variant intronic to *LRP2* (rs12988804), previously reported to be associated with relapse risk,^23^ and annualised relapse rate (ARR). However, we were unable to find an association between this variant and ARR in our targeted analysis (p = 0.925).

## Discussion

Two of the fundamental, unanswered questions with respect to relapsing-remitting MS are first, what is the source of the marked clinical disease heterogeneity? That is, why do some people with RMS have a rapidly progressing, severely disabling disease course, whilst others do not? And second, can we utilise genetic and other information to predict MS outcomes?

Here, through a series of analyses that took advantage of a unique, multicentre, prospectively ascertained, longitudinal, clinical dataset,^24^ we can shed some light on the genetic architecture that underpins MS clinical heterogeneity. Our primary, unbiased GWAS analyses demonstrate that there are no common variants with moderate to large effect sizes that contribute to MS severity. With time, and very large cohorts, we will likely confirm that MS severity is at least partially determined by polygenic mechanisms of small effect size. Alternatively, we may find that variants which influence severity may be time-variable, rather than having a constant effect.^25^ Importantly, our results suggest that disease outcomes are not under strong genetic control. Indeed our study results demonstrated that common genetic variants explained only 20% of severity heritability, with wide error margins. Therefore, suggesting that, as clinical experience shows, outcomes are, to an extent, modifiable with appropriate and early disease-modifying therapy (DMT) intervention.^26-28^ This is further underscored in the modern era where, with the introduction of DMT, rates of disability accumulation have slowed, and fewer disabling cases are being seen, relative to historical cohorts.^29-33^ Future pharmacogenomic studies^34^ may prove to be invaluable to guide precise prescription practices to further slow progression or modify disease outcomes.

The complex interplay between genes and the environment likely additionally plays a significant role in outcome modulation. It has been shown that disability accumulation may be modified by additional lifestyle factors such as pregnancy^27^ and smoking cessation.^35^ Epigenetic studies may therefore shed further light on relevant, modifiable mechanisms that regulate MS outcomes.

The application of machine learning to GWAS data is considered by some^36^ to be the “last hope” to gain meaningful insights for complex diseases where no variants meet significance thresholds.

Our machine learning algorithm was unable to provide additional biological insights into the underlying genetic architecture of MS severity, instead reinforcing that common SNVs independently contribute miniscule weights towards determining MS severity. Regardless, machine learning was able to predict non-linear effects and large SNV clusters that can accurately classify MS outcomes, and may prove to be of prognostic utility. Whilst we retained the MS susceptibility wGRS in our algorithm, it was not a major contributor to our predictive model, again consistent with our above findings, and past reports demonstrating that MS risk variants have little influence on severity outcomes.^7^ The classification accuracy of our machine learning algorithm increased with the addition of age at onset. In fact, age at onset was one of the strongest predictors of outcome in our machine learning models, consistent with past reports.^27^ Our machine learning algorithm was designed with internal checks to prevent data over fitting. We used a slow learning rate, with a 70/30 training/testing set and internal bootstrapping over 20,000 learning iterations. We achieved positive predictive values for outcome assignation of between 0.889 – 0.844 and negative predictive values in the range of 0.851 – 0.853. To our knowledge, this is a world first for MS genetic studies. Whilst a previous ML study successfully predicted MS severity,^37^ this was predicated on health records, and data that take years to decades to obtain e.g. change in clinical parameters between years ‘x’ and ‘y’ to predict ‘z’. The variables included in our classification algorithm are readily available at disease onset. With the rapid decrease in the cost of beadchip genotyping, and high PPV and NPV we achieved, our machine learning algorithm could readily translate into clinical practice upon validation in an independent cohort.

In our secondary analyses, we replicated the association between the main MS risk allele HLA-DRB1*15:01 and age at onset.^4, 6^ Further, ours is the first study to replicate rs868824, intronic to *IMMP2L*,^10^ as being associated with age at onset in a targeted analysis. *IMMP2L*, an inner mitochondrial membrane protease, has been associated with cellular senescence,^38^ ovarian aging via oxidative stress and estrogen-mediated pathways,^39^ together with neurological disorders.^40-43^ Recent evidence points to accelerated cellular senescence and biological aging in people MS,^44-46^ and suggests that these factors may reduce remyelination capacity.^45^ The validation of the association of rs868824 with age at onset, is a first step towards understanding the potential biological mechanisms underlying accelerated cellular senescence in MS.

The integration of the top SNVs identified in our *de novo* GWAS analyses into hard EDSS disability milestone survival analyses, again identified variants intronic to or near genes implicated in mitochondrial function: rs111399831, nearest to *SUCLA2*, and rs9643199 intronic to *MTSS1*; as well as variants intronic to genes implicated in CNS function: rs7289446 intronic to *SEZ6L*, rs295254 intronic to *RCL1*, rs9643199 intronic to *MTSS1*, rs2776741 nearest to *RCAN3AS*, and rs7070182 Intronic to *TCF7L2*. The latter three having sex-specific effects. The hazard ratios associated with reaching irreversible EDSS 3 or 6, conferred by carriage of the minor allele at each SNV, were consistent with the effect sizes of these variants in our GWAS analyses: that is, effect sizes were small, but significant in survival analyses. Most importantly, these variants identify highly biologically and clinically plausible leads for potential replication of clinical heterogeneity in similar or larger cohorts.

*SUCLA2* is expressed in the brain and muscle, and encodes the beta-subunit of succinate-CoA ligase, an enzyme required for the maintenance of mitochondrial DNA.^47^ Variations in *MTSS1* have been reported to associate with mitochondrial complex 1 deficiency in ClinVar (SCV001137705.1, SCV001137706.1). The variants we describe here are intronic to, or near to these genes, and are likely to be tagging rather than causal. However, together with *IMMP2L*, we describe three variants associated with mitochondrial function, where mitochondrial dysfunction is a recognised pathophysiological hallmark of CNS injury in MS.^48, 49^

Whilst *MTSS1* has been reported to associate with mitochondrial function, it has primarily been described in the context of B-cell mediated immunity,^50^ and various CNS pathologies.^51,52^ Most relevant perhaps to MS outcomes, is the association between *MTSS1*, cortical volume, and Purkinje cell dendritic arborization.^53, 54^ *SEZ6L*, the top signal in both l-ARMSS and l-MSSS analyses, is implicated in dendritic spine density variation, and arborization in the hippocampus and somatosensory cortex.^20^ Disruptions in *SEZ6L* cause neurodevelopmental, psychiatric, and neurodegenerative conditions, as well as having a role in motor function.^20, 55, 56^ Copy number variation in *RCL1*, has also been associated with severe psychiatric disease,^57^ and depression.^58^ Progressive synaptic loss, or synaptopathy, is a hallmark of MS pathology;^59, 60^ evident in both acutely active demyelinating lesions,^61^ as well as chronic inactive lesions.^62^ It has been shown that loss of synaptic density is associated with network dysfunction,^60^ implicating a failure of synaptic plasticity to compensate for immune-mediated neural damage. It is therefore plausible that the variants identified in this study implicate a genetic susceptibility to impaired compensatory mechanisms, or impaired neural survival in those with severe MS. This of course requires independent validation but raises an intriguing new line of enquiry.

Interestingly, we identified a variant intronic to *TCF7L2* as associated with severity in males. *TCF7L2* is a transcription factor involved in Wnt signalling pathways, and associated with neurodevelopmental disorders.^63^ Critically in the context of our study, *TCF7L2* has been shown to maintain oligodendrocyte progenitor cells in the progenitor state, acting as a molecular switch that can inhibit Wnt signalling to promote oligodendroglial differentiation.^64^ Why this variant was associated with severity only in males in our study is unclear. However, the role of *TCF7L2* in MS severity requires further investigation. Interestingly, gene set enrichment analyses revealed that both Wnt signalling pathway components together with progesterone response pathways were enriched in our analyses. Progesterone is a known regulator of myelin development, as well as having neuroprotective effects,^65^ lending support to the notion that genetic susceptibility to impaired remyelination predisposes to more severe MS.

Our tissue enrichment analyses specifically pointed towards genes enriched in cerebellar function. The cerebellum plays a key role in motor coordination as well as cognition.^66^ It has long been held^26^ and recently confirmed,^67^ that cerebellar signs and symptoms are a predictor of poor prognosis in MS. The results of our analyses therefore point to highly relevant and biologically plausible genetic explanations for clinically observed disease heterogeneity.

Our multicentre study was conducted using rigorously defined and prospectively collected longitudinal clinical and treatment data from the MSBase Registry, making our cohort globally unique. Due to the nature of this cohort, we were unable to validate our results in an equivalent dataset, therefore, the data presented herein require independent validation. We did try to overcome this limitation by testing top SNV signals using alternate definitions of disease severity, namely survival analyses of time to irreversible disability milestones. Similarly, our ML analyses were performed using a conservative 70/30 training/testing split relative to the traditional 80/20 split, accompanied with internal bootstrapping. Our efforts to expand our cohort for future analyses are ongoing.

Here we report an important milestone in our progress towards understanding the clinical heterogeneity of MS outcomes, implicating functionally distinct mechanisms to MS risk. We demonstrate that common genetic variants of moderate to large effect sizes do not contribute to MS severity. In secondary sex-stratified analyses, we identified two genetic loci that surpassed GWAS significance thresholds, providing evidence of sex dimorphism in MS severity. We identified a further six variants with strong evidence for regulating clinical outcomes. We observed an overrepresentation of genes expressed in CNS compartments generally, and specifically in the cerebellum. These involved mitochondrial function, synaptic plasticity, cellular senescence, calcium and g-protein receptor signalling pathways. Importantly, we demonstrate that machine learning using common SNV clusters, together with clinical variables readily available at diagnosis can improve prognostic capabilities at diagnosis, that which goes beyond T2 MRI lesion load,^68^ and with further validation has the potential to translate to meaningful clinical practice change.

## Methods

### Study population

Participants were recruited from eight tertiary-referral MS-specialist centres, from 3 countries (Australia, Spain, and Czech Republic), participating in the MSBase Registry. MSBase is an international, prospective, observational, MS clinical outcomes registry, registered with the World Health Organization International Clinical Trials Registry Platform, ID ACTRN12605000455662.^24^ Data are entered by neurologists in, or near real-time including: participant demographics, disease phenotype, expanded disability status scale (EDSS) scores, relapse information, and disease modifying therapy use. Clinical assessments occur on average every 6 months.

### Ethics approvals

This study was approved by the Melbourne Health Human Research Ethics Committee, and by institutional review boards at all participating centres. All participants gave written informed consent for participation in the MSBase Registry, together with additional informed consent to participate in genetic research (HREC/13/MH/189 and per local approvals elsewhere).

### Study Inclusion Criteria

People with MS (pwMS) of European ancestry with clinically definite, relapse-onset MS, based on McDonald criteria^69-71^ and participating in MSBase. Further, minimum inclusion criteria comprised: sex, birthdate, age at onset, ≥5 years of symptom duration; ≥5 years prospective follow-up in MSBase; ≥3 EDSS scores recorded in the absence of a relapse (defined as EDSS scores recorded within 30 days of relapse onset date); availability of relapse and treatment history. Symptom duration was calculated based on the most recently recorded EDSS visit.

### Phenotyping, severity assignation and recruitment

Data used for phenotyping pwMS were extracted from the registry on 4 September 2019. EDSS scores recorded in the absence of a relapse were used to calculate an age-related MS severity (ARMSS)^19^ score and MS Severity Scale^13^ (MSSS) score. It has been demonstrated that at the individual level, that cross-sectional MSSS cannot be used for prognostication,^13^ but that longitudinal MSSS scores may be less noisy in individual prognostics.^72^ Therefore, for each participant, we calculated the median longitudinal ARMSS (l-ARMSS) and median longitudinal MSSS (l-MSSS) using each available ARMSS or MSSS score (minimum 3 scores, Tables 1 & S2). Median relapse-independent l-ARMSS and l-MSSS scores were then divided into quintiles to stratify the cohort for severity. The top and bottom quintiles were defined as outcome extremes. Our definitions of mild and severe disease were based on all individuals meeting minimum inclusion criteria (n=5,851; Figure S1). Participant recruitment was then enriched for those at outcome extremes. Lists of study participants in the top and bottom decile of severity were sent to centre PIs to ensure accurate diagnosis. In cases of diagnostic uncertainty, or re-classification (e.g. ADEM, primary progressive MS), these pwMS were excluded from our study. A further age at onset (AAO) phenotype was defined as age at first symptom onset.

Symptom duration was defined as the number of years between first symptom onset and the most recently recorded clinical visit reported by a neurologist in MSBase. Follow-up was defined as the number of years between first neurologist recorded visit in MSBase, and the most recent clinical visit. Percentage of time exposed to disease-modifying therapy (DMT) was defined as the total time exposed to any approved MS DMT as a percentage of symptom duration, as recorded in MSBase.

### Statistical analyses

Data processing and statistical analyses were performed in Stata v17 (Stata Corp, College Station, TX) or R (http://R-project.org). Monash high performance computing infrastructure through MASSIVE was used for big data manipulation and computationally extensive analyses.^73^ Continuous variables were assessed for normality using the Shapiro-Wilk normality test, and described as mean and standard deviation (SD) or median with interquartile range (IQR), as appropriate. Categorical variables were described using frequencies. Standardised differences between key demographic and clinical variables were assessed using the Cohen’s d statistic. Correlations between l-ARMSS and AAO and the weighted genetic risk score (wGRS) were assessed using Pearson’s correlation coefficients. All analyses were 2-tailed.

### Genotyping, imputation and quality control

Detailed methodology can be found in supplementary materials and methods. Briefly, whole blood gDNA was genotyped using the Illumina MegaEx BeadChip array. This array was customized with an additional 3,000 single nucleotide variants (SNVs) of interest including: known MS risk SNVs, a suite of tag SNVs to classical HLA alleles,^11^ previously published putative severity SNVs,^10, 14, 15, 17^ and others of interest, including SNVs previously associated with neurodegeneration in other diseases (see supplementary materials). Samples were genotyped in two tranches. Each tranche containing DNA from Australia, the Czech Republic and Spain. Genome-wide data was therefore organised into six data sets. Following rigorous per-data-set quality control (supplementary methods, Table S1), we imputed all samples using the Haplotype Reference Consortium panel (r1.1)^74^ on the Michigan imputation server, resulting in 22,469,259 SNVs. After further per-SNP QC, this resulted in 5,985,626 (final) SNVs with a minor allele frequency of at least 5%.

### Association testing

PLINK (v 1.9 and v 2.0)^75^ were used to conduct association testing and meta-analyses. For continuous traits, we used linear regression to analyse each of the 6 data sets, adjusted for the first 5 principal components (PCs), weighted genetic risk scores^76^, disease-modifying therapy use, together with variables identified to have a standardised difference greater than 15% between severity extremes. Combined data set results of all 6 groups were then analysed using fixed-effects meta-analyses to identify statistically independent SNVs.

For binary traits, all 6 groups were analysed jointly, due to lower sample numbers. We used logistic regression, adjusting for group ID, the first 5 PCs together with covariates with Cohen’s d >0.15. For replication analyses, the Bonferroni-deflated p-value to meet replication threshold was set to p ≤ 4.31×10^−4^ (0.05/116 replication SNVs). The *de novo* genome-wide association study (GWAS) p-value threshold was set to p<5×10^−8^. SNVs meeting 1×10^−8^< p <5×10^−5^ were considered to have nominal evidence of association with the trait of interest. Weighted genetic risk scores (wGRS)^76^ were calculated based on directly genotyped SNVs described by the IMSGC (supplementary material).^3^ Calculations estimated a sample size of 915 individuals per group was required to achieve 80% power to detect an SNV with MAF 0.2 and an odds ratio of 1.3, based on binary severity outcomes.

### Survival analyses

We assessed time to reach the hard disability milestones of irreversible EDSS 3 and 6 for those individuals who had not yet reached these at first MSBase-recorded clinic entry. Where disability milestones were not met during study observation, data were censored at the most recent clinic visit. Survival analyses were based on carriage of the minor allele for SNVs at the top 10 nearest genes identified in our l-ARMSS and l-MSSS *de novo* association analyses; and top 5 nearest genes identified in our sex-stratified l-ARMSS and l-MSSS analyses. Cox proportional hazards regression (implemented in Stata v17) was used to calculate hazard ratios (HRs) with 95% confidence intervals (CI). Multivariable models were adjusted for AAO, MS susceptibility wGRS, percentage of follow-up exposed to DMT and sex. The Schoenfeld residuals global test was used to detect a violation of the Cox proportional hazards assumption. Where the proportionality assumption was violated, a Weibull regression approach was applied. Survival data were visualised using Kaplan-Meier curves.

### Heritability analysis

Narrow sense heritability (h_snp_) was estimated from individual-level GWAS data using a genome-based restricted maximum likelihood (GREML) approach implemented using GCTA software,^77^ and BOLT-LMM.^78^ BOLT-LMM was additionally used to estimate per-chromosome heritability estimates. We further employed a summary statistic approach using a linkage disequilibrium (LD) score regression (LDSC) implemented in LDSC software.^79^

### Enrichment analyses

We used FUMA^22^ to assign SNVs (p <1×10^−5^) to Genes, Tissues and functions using as per online instructions. The Panther database^80^ was used to further confirm gene pathways/ontologies that were over-represented and enriched in the variants with top association hits (p <1×10^−5^). Tissue expression of variants with p<1×10^−05^ was further validated using both the TissueEnrich package^81^ and Geneanalytics database^82^.

### Machine Learning

We chose to implement non-linear machine learning (ML) models for severity classification as linear ML models, that do not account for interaction between genetic variants, have been found to perform no better than simple linear regression in the context of common variant-based disorders.^83^ All SNVs that had a p-value of 0.01 or less in the *de novo* meta-analyses were used to generate datasets compatible with gradient boosting algorithms (xgboost^21^). A total of 62,351 SNVs were included, with binary l-ARMSS score severity as the outcome. A training set of 70% of the pwMS was randomly selected, ensuring a balanced representation of severe and mild MS outcomes. After training with internal bootstrapping of 0.7 for 10k iterations, the model was tested on the 30% of the remaining cohort, i.e. those datasets never encountered by the algorithm. We were cautious to avoid overfitting our models by using 70%-30% cut off for the train/test data sets; a more conservative approach than others that tend to use a 80%-20% cut off.^84^ Further, a slow learning rate (eta = 0.01) was implemented to avoid overfitting.^85, 86^ The algorithm calculated a prediction score for each new individual regarding their severity group membership. Accuracy of prediction was compared to the clinically-informed grouping of each individual.

Using the prediction values generated on the test set for each model, as well as the true membership values of each sample, a confusion matrix was generated along with accuracy, sensitivity, specificity, and Kappa statistics using the confusion matrix function of Caret package. Furthermore, to evaluate the prediction accuracy and performance of the models, the Receiver Operator Characteristic (ROC) curve was plotted to explore the relationship between false positives and negatives, and the Area Under the Curve (AUC) for each model was calculated.

## Data Availability

Clinical data from the MSBase Registry: To protect participant confidentiality, de-identified patient-level data sharing may be possible in principle, but will require permissions/consent from each contributing data controller.
Genetic data will be deposited to an access controlled database shortly, whilst we continue to explore these data in further analyses. Access requests with scientifically sound proposals can be made in writing to Dr Vilija Jokubaitis or Prof Helmut Butzkueven.

## Acknowledgements

We thank all the people with MS who participated in this research without whom this work would not be possible. We would also like to acknowledge the patients and the Biobank Nodo Hospital Virgen Macarena (Biobanco del Sistema Sanitario Público de Andalucía) integrated in the Spanish National biobanks Network (PT20/00069) supported by ISCIII and FEDER funds, for their collaboration in this work.

The authors would like to acknowledge Prof David Booth, research nurses Ms Jo Baker, Ms Jodi Haartsen, Ms Sandra Williams, Ms Lisa Taylor for assisting with sample collection for this study, and Ms Malgorzata Krupa for research assistance. Further, the authors acknowledge the Center for Genome Technology within the University of Miami John P. Hussman Institute for Human Genomics for generating all the MegaEx array genotype data for this project and specifically Anna Konidari for overseeing the genotyping efforts and assistance with the Illumina custom design process.

This work was supported by a Research Fellowship awarded to Dr Vilija Jokubaitis from Multiple Sclerosis Research Australia (16-0206), and research grant support from the Royal Melbourne Hospital Home Lottery Grant (MH2013-055), Charity Works for MS (2012 Project grant), MSBase Foundation Project Grant, and Monash University. EH, DH, PK are supported by the Czech Ministry of Education, project PROGRES Q27/LF1. FM and AA receive support from the Agencia Española de Investigación (AEI)-Fondos Europeos de Desarrollo Regional (FEDER) (PID2019-110487R-C21) and Junta de Andalucía (P18-RT-2623).

## Author contributions

**VGJ** conceived and designed the study, obtained data, performed data analysis, interpreted the data, and drafted the manuscript.

**OI** contributed to study design, performed data analysis, interpreted the data, and substantively revised the manuscript

**JS** contributed to study design, performed data analysis, interpreted the data, and substantively revised the manuscript

**PK** obtained study data, interpreted the data, and substantively revised the manuscript

**FM** contributed to study design, obtained study data, and substantively revised the manuscript

**DH** performed data analysis, and substantively revised the manuscript

**SE** obtained study data, interpreted the data, and substantively revised the manuscript

**JLS** obtained study data, interpreted the data, and substantively revised the manuscript

**MS** obtained study data, interpreted the data, and substantively revised the manuscript

**RL** interpreted the data, and substantively revised the manuscript

**TJK** obtained study data, interpreted the data, and substantively revised the manuscript

**TK** obtained study data, and substantively revised the manuscript

**PDJ** obtained study data, and substantively revised the manuscript

**AB** contributed to data analysis, and substantively revised the manuscript

**JLM** contributed to data analysis, and substantively revised the manuscript

**BVT** obtained study data, interpreted the data, and substantively revised the manuscript

**SV** obtained study data, and substantively revised the manuscript

**LL** contributed to study analysis, and substantively revised the manuscript

**KV** obtained study data, and revised the manuscript

**MIGS** obtained study data, and revised the manuscript

**AA** obtained study data, and revised the manuscript

**AvdW** obtained study data, and substantively revised the manuscript

**EKH** obtained study data, and substantively revised the manuscript

**GI** obtained study data, interpreted the data, and substantively revised the manuscript

**NP** contributed to study design, performed data analysis, interpreted the data, and substantively revised the manuscript

**DH** obtained study data, interpreted the data, and substantively revised the manuscript

**HB** conceived and designed the study, obtained data, interpreted the data, and drafted the manuscript.

## Competing interests

The authors report no competing interests

## Notes

### Competing Interest Statement

The authors have declared no competing interest.

### Funding Statement

This work was supported by a Research Fellowship awarded to Dr Vilija Jokubaitis from Multiple Sclerosis Australia (16-0206), and research grant support from the Royal Melbourne Hospital Home Lottery Grant (MH2013-055), Charity Works for MS (2012 Project grant), MSBase Foundation Project Grant, and Monash University.

