## Supplemental Methods, Materials and Figures for "Not all roads lead to the immune system: The Genetic Basis of Multiple Sclerosis Severity Implicates Central Nervous System and Mitochondrial Involvement"

**This PDF file includes:**

Materials and methods

Figs S1 to 12

References

**Other supplementary material for this manuscript includes the following:**

Table S1 to S22 as separate Excel file: Supplementary_Tables_Jokubaitis_et_al.xlsx

Please note, the names of supplementary tables and figures in this document replicate the order in the main manuscript.

### *Supplementary Materials and Methods*

#### *Cohorts:*

Participants were recruited from eight tertiary-referral MS-specialist centres, from 3 countries (Australia, Spain, and Czech Republic), participating in the MSBase Registry.[1] Australian participants were recruited from the Royal Melbourne Hospital, Melbourne, Victoria (n=305); The Box Hill Hospital, Melbourne, Victoria (n=225), The Flinders Medical Centre, Adelaide, South Australia (n=96), The John Hunter Hospital, Newcastle, New South Wales (n=88), The Royal Hobart Hospital, Hobart, Tasmania (n=12), and Westmead Hospital, Sydney, New South Wales (n=3). The Czech cohort was recruited from the General University Hospital, Prague (n=761). The Spanish cohort was recruited from Hospital Universitario Virgen Macarena, Sevilla (n=494). Cohort characteristics, and the samples that passed QC are detailed in supplementary Tables S1 - S4.

Individuals contributing to the extremes of l-ARMSS and l-MSSS outcomes binary phenotypes were not identical. A total of 1239 (68.3%) of the cohort were classified as being at the extremes of outcome by either l-ARMSS or l-MSSS metrics. 346 individuals overlapped contributing to the mild phenotype across both l-ARMSS and l-MSSS, 50.4% of all those classified as mild by either phenotype. Conversely, 377 individuals overlapped contributing to the severe phenotype across both l-ARMSS and l-MSSS, 68.2% of all those classified as severe by either phenotype (TableS2).

Samples were genotyped across two tranches, as sufficient samples were collected at each site. Each tranche contained samples from all 3 countries.

Total included patients are outlined in Figure S2.

#### *MegaEx array custom content*

The Illumina MegaEx BeadChip array was used as our platform genotyping array. We added 3K custom single nucleotide variants (SNVs) of interest designed using the Illumina iSelect platform. These included: known MS risk SNVs based on the IMSGC 2013 GWAS,[2] a suite of tag SNVs to classical HLA alleles,[3] previously published putative severity SNVs,[4-7] and others of interest, including SNVs previously associated with neurodegeneration in other diseases. The custom SNV content is found in the file S1: “SNV list-JokubaitisMSGPstudy”.

#### *Quality Control:*

The same quality control (QC) pipeline for the 22 autosomal chromosomes was applied to each of the 6 data sets. For excluded samples and SNVs see supplementary Table S1.

Single Nucleotide Variants (SNVs) were excluded based on low call rate (<95%), low minor allele frequency (MAF < 0.05), violation of Hardy–Weinberg equilibrium ($p < 1\times10^{-5}$), monomorphism and non-autosomal location.

Samples were excluded based on sex inconsistencies and low call rate (<95%). We then LD-pruned the autosomes in *PLINKv1.9* using indep-pairwise 100 2 0.1 (100 SNVs window, 2 SNVs step, r2 = 0.1). The LD-pruned datasets were then used to exclude individuals with an inbreeding coefficient F≥0.05 or ≤-0.05. Relatedness was assessed using Identity by Descent (IBD) analysis in *PLINKv1.9* and those with pi-hat > 0.1, corresponding to 3rd and 4th degree relatives, were excluded. IBD was confirmed using *KING.*[31] Principal components (PC) analysis was implemented in *EIGENSTRAT*[32] and individuals that were outside ± 6 standard deviations of the each of the first 10 PCs were excluded. PCs were projected to HapMapIII data to assess population stratification effects (Figure S1a-f), and we manually excluded population outliers falling outside the main European ancestry cluster.

#### *IMPUTATION:*

Genotype VCF files were prepared for imputation as per instructions on the Michigan Imputation Server website (https://imputationserver.readthedocs.io/en/latest/prepare-your-data/). Genotypes were then imputed using Haplotype Reference Consortium (HRC) version r1.1[8] (https://imputationserver.sph.umich.edu/index.html#!), and converted to genotype calls in *PLINKv1.9*. Again, imputed SNVs with low minor allele frequency (MAF < 0.05), those in violation of Hardy–Weinberg equilibrium ($p < 1\times10^{-5}$), monomorphism and non-autosomal location were excluded from the analysis.

#### *WEIGHTED GENOMIC RISK SCORE (wGRS):*

Of 200 non-HLA MS-associated SNVs reported by the IMSGC[9], 198 were SNV genotyped in the current study and included in the calculation of our wGRS. Of the 198 SNVs, 195 were found with minor allele count (MAC) of 20 or more across the 6 cohorts, hence were included in the meta-analysis. The p-values and beta values for these 195 SNVs are reported in Table s5. The number of minor alleles (0, 1, 2) at each of the 198 loci was multiplied by the log odds ratio reported by the IMSGC,[9] implemented in R (version 4.1.1). This wGRS was then included in adjusted *de novo* association analyses.

#### *HERITABILITY ESTIMATES:*

Narrow sense heritability ($h_2g$) results are detailed in table S18. We used 3 different tools (GCTA, LDSC, and BOLT-LMM) to assess and corroborate our findings.

Below are the different parameters used with each package. For each model we used 4942008SNVs with MAF > 0.01 and 1813 samples.

1- Using GCTA, we created variance components (genetic relationship matrices; grms) for:

A) Genome Wide SNVs to estimate overall heritability used the original genotypes that passed QC (4942008) and the respective phenotype file (ARMSS or MSSS) used in the meta-analysis.

B) Per-chromosome SNVs to estimate grm for each chromosome. GCTA multi-GRM combined the results of all chromosomes for each trait (ARMSS and MSSS).

2- BOLT-LMM was used with the same genotype and phenotype files used for GCTA for each trait.

3- LDSC package used the summary statistics from the original meta-analysis file for each phenotype, and calculated variance from European reference population files recommended by LDSC package. The LD reference panel contained 1290028 SNVs, and the final LDSC model used 1067715 SNVs.

#### *ENRICHMENT ANALYSES:*

To identify the genes nearest to the identified SNVs ($p<1x10^{-05}$), to perform tissue enrichment and gene set enrichment analyses, we uploaded GWAS results for each of the l-ARMSS, l-MSSS and AAO phenotypes into FUMA.[10] Results were corroborated using PANTHER pathways tool.[11, 12]

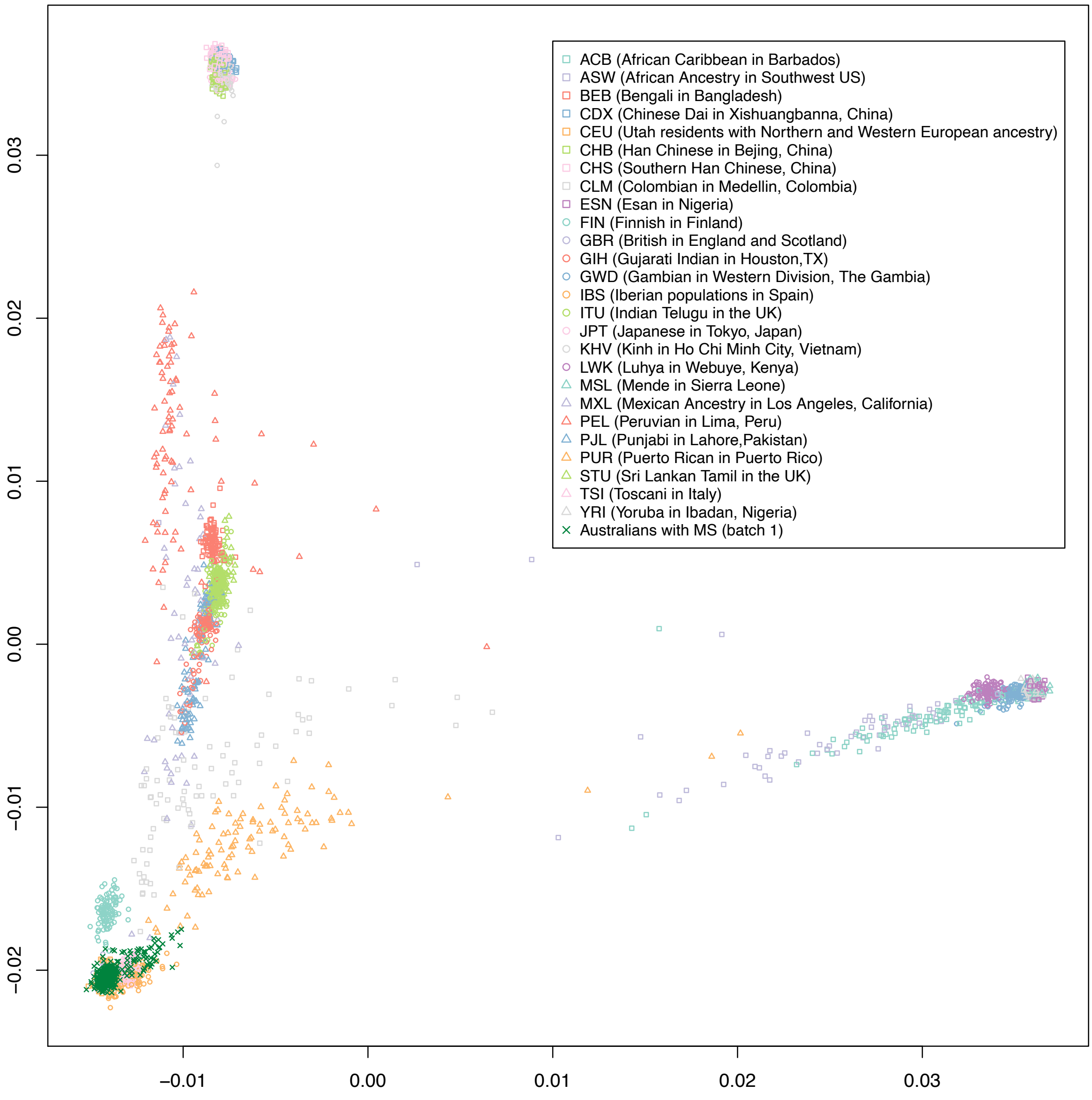

***Figure S1a:*** Australian cohort (Batch 1) PCs projected to HapMapIII data shows clustering with European populations.

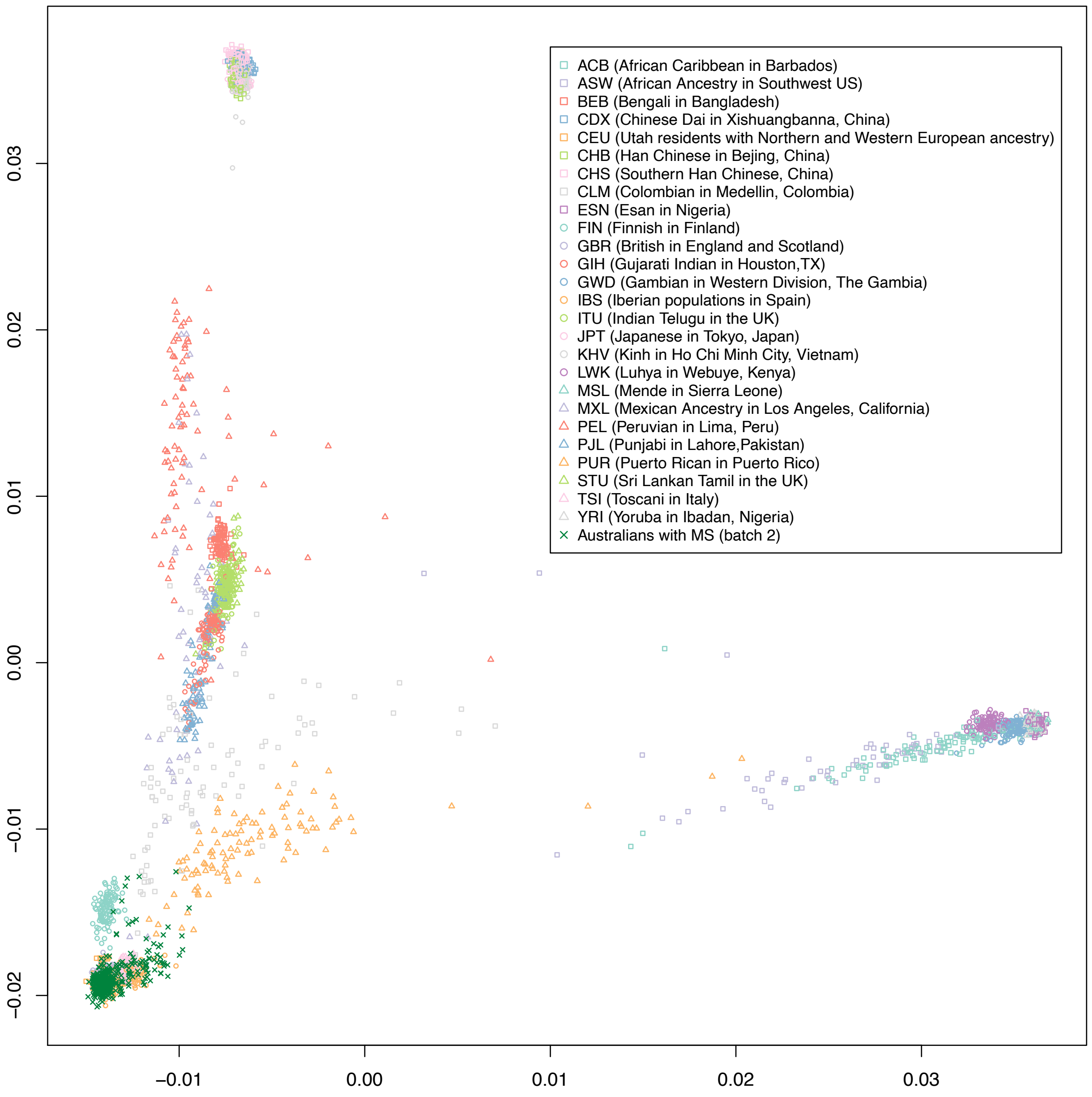

***Figure S1b:*** Australian cohort (Batch 2) PCs projected to HapMapIII data shows clustering with European populations.

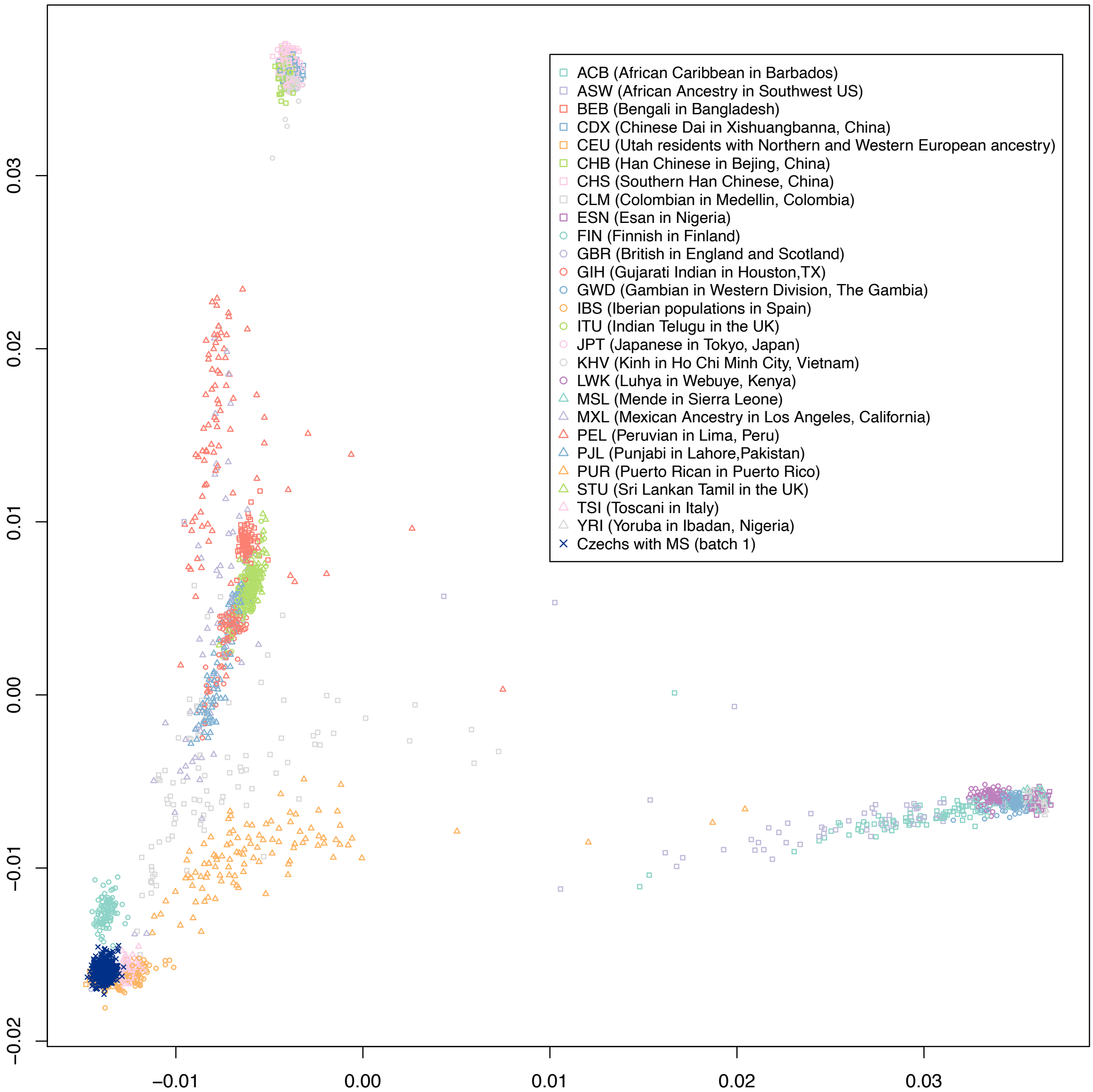

***Figure S1c:*** Czech Republic cohort (Batch 1) PCs projected to HapMapIII data shows clustering with European populations.

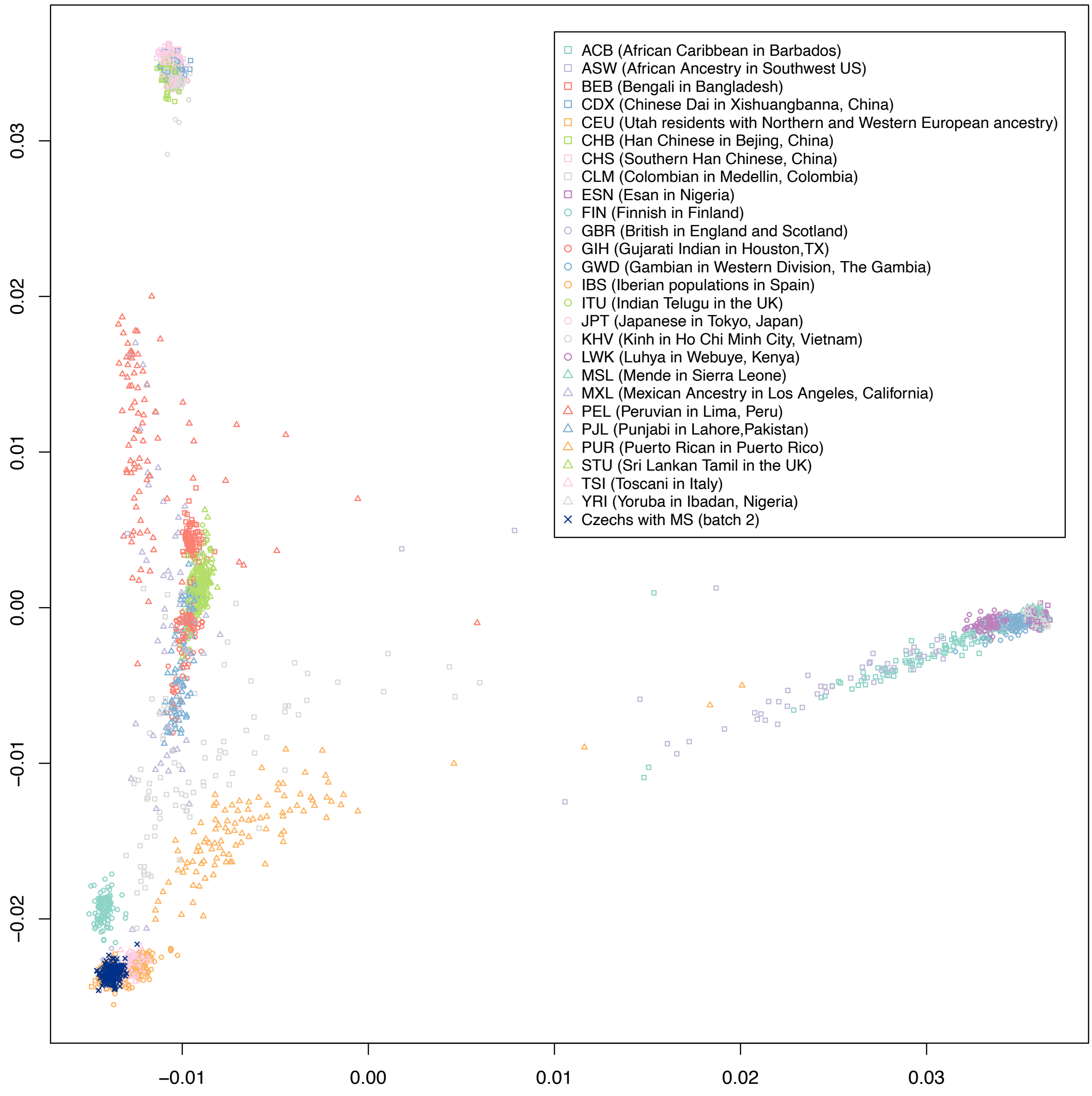

***Figure S1d:*** Czech Republic cohort (Batch 2) PCs projected to HapMapIII data shows clustering with European populations.

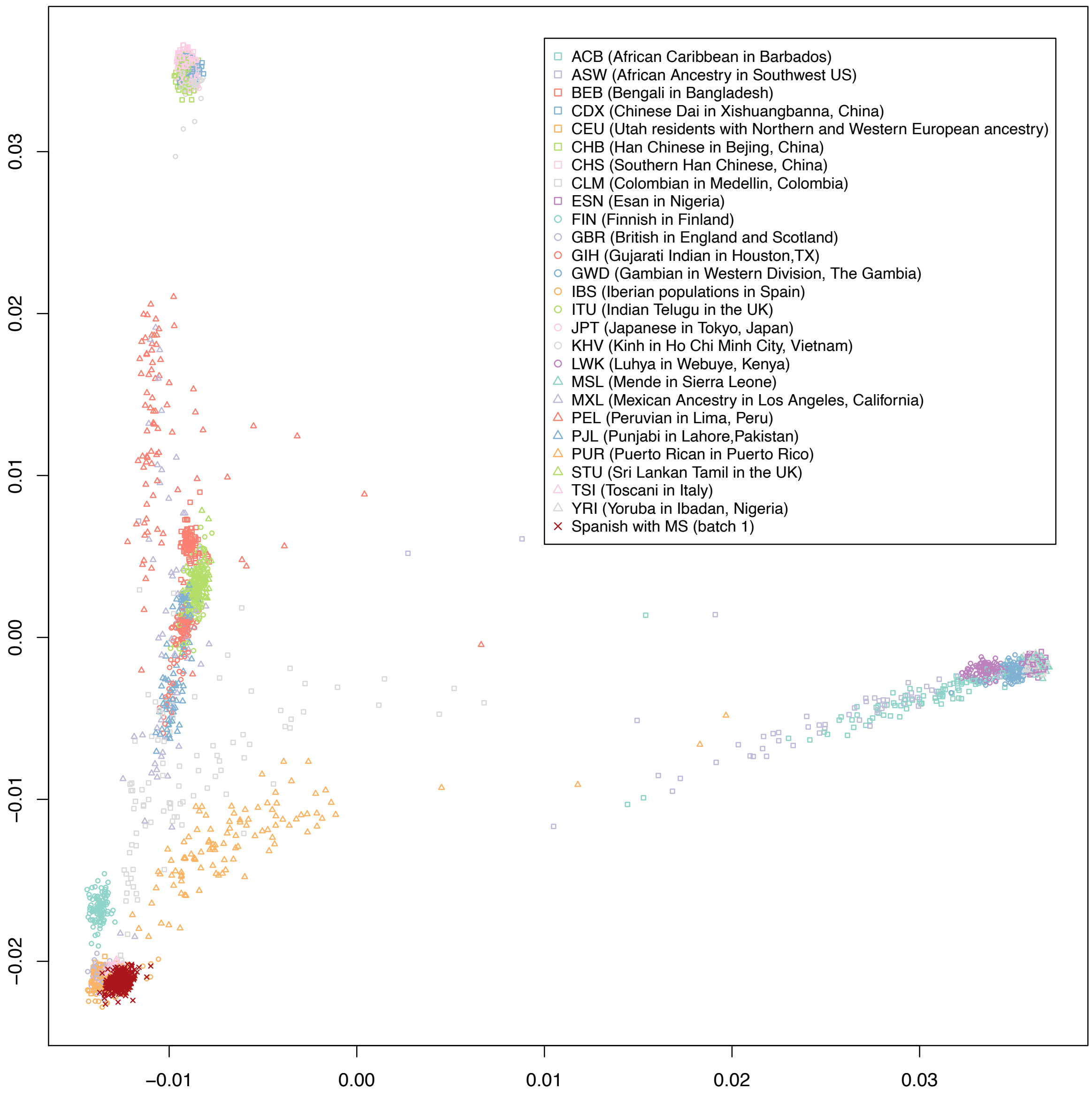

***Figure S1e:*** Spanish cohort (Batch 2) PCs projected to HapMapIII data shows clustering with European populations.

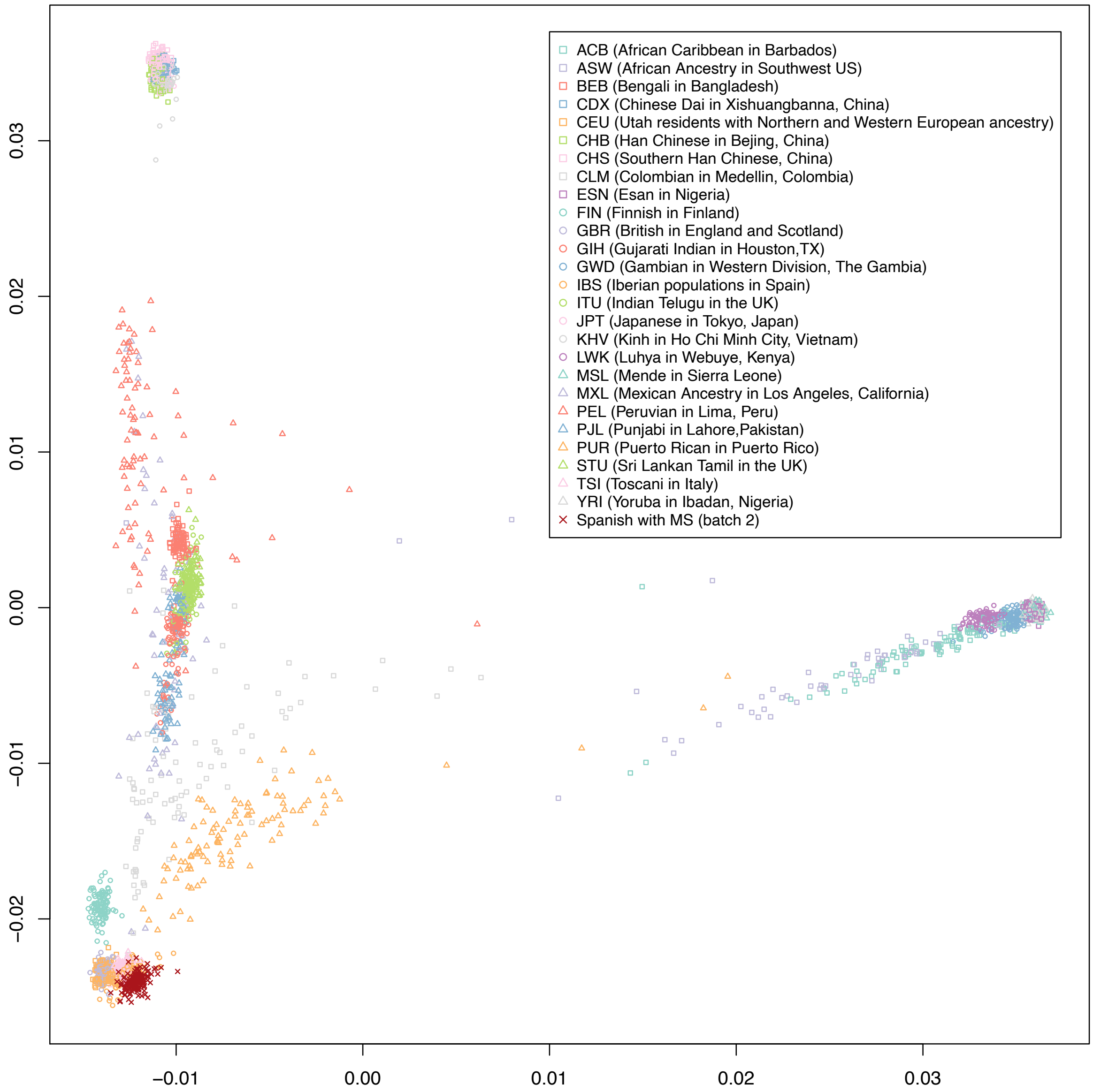

***Figure S1f:*** Spanish cohort (Batch 2) PCs projected to HapMapIII data shows clustering with European populations.

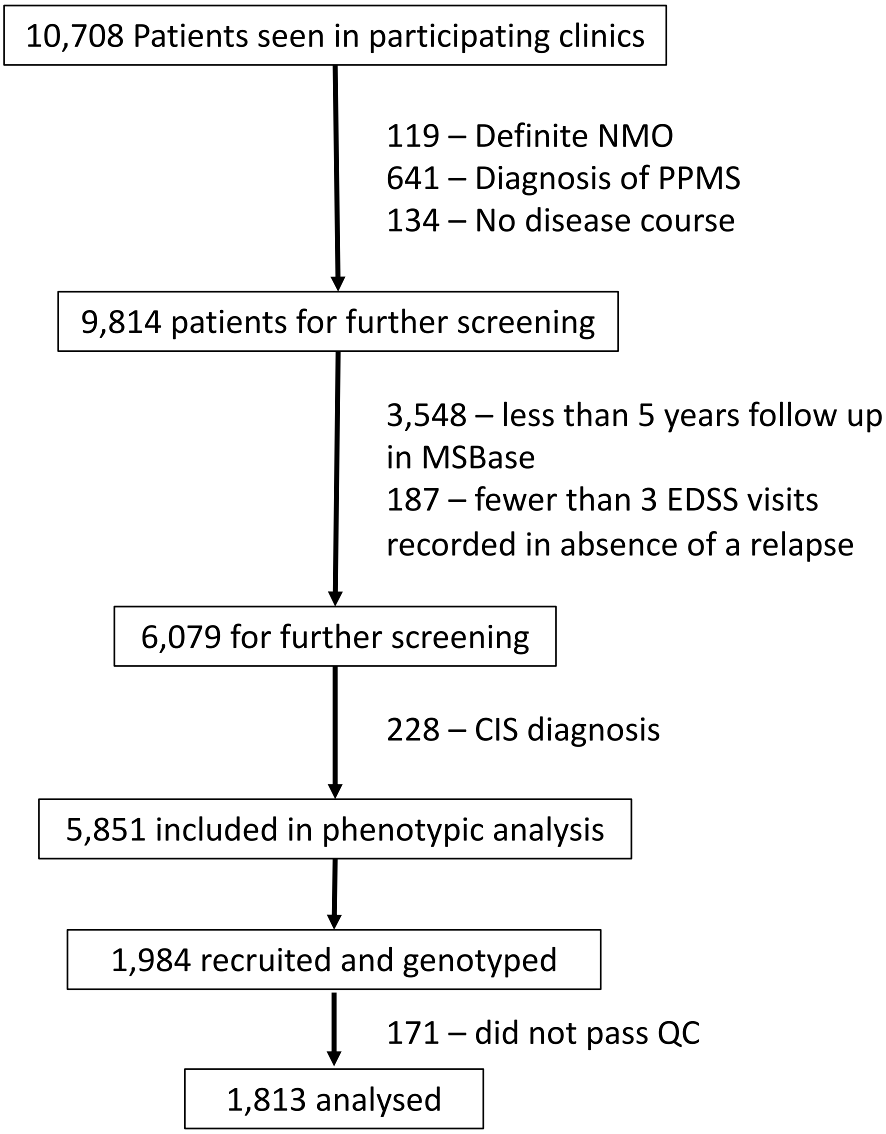

***Figure S2:*** Summary of patients screened for inclusion and analysis

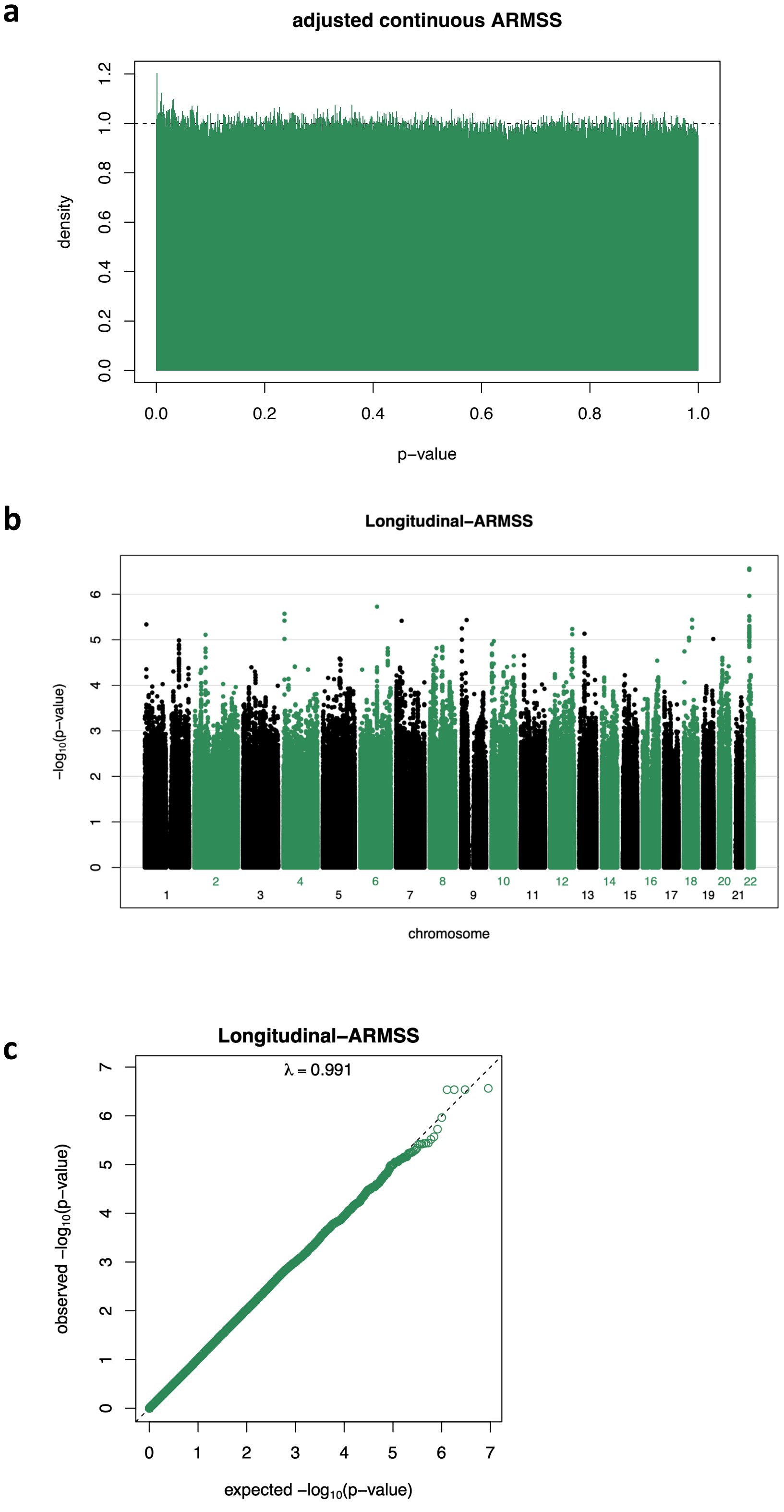

***Figure S3:*** Continuous l-ARMSS GWAS adjusted fixed-effects meta-analysis **a:** p-value distribution **b:** Manhattan plot **c:** QQ plot and genomic inflation factor λ

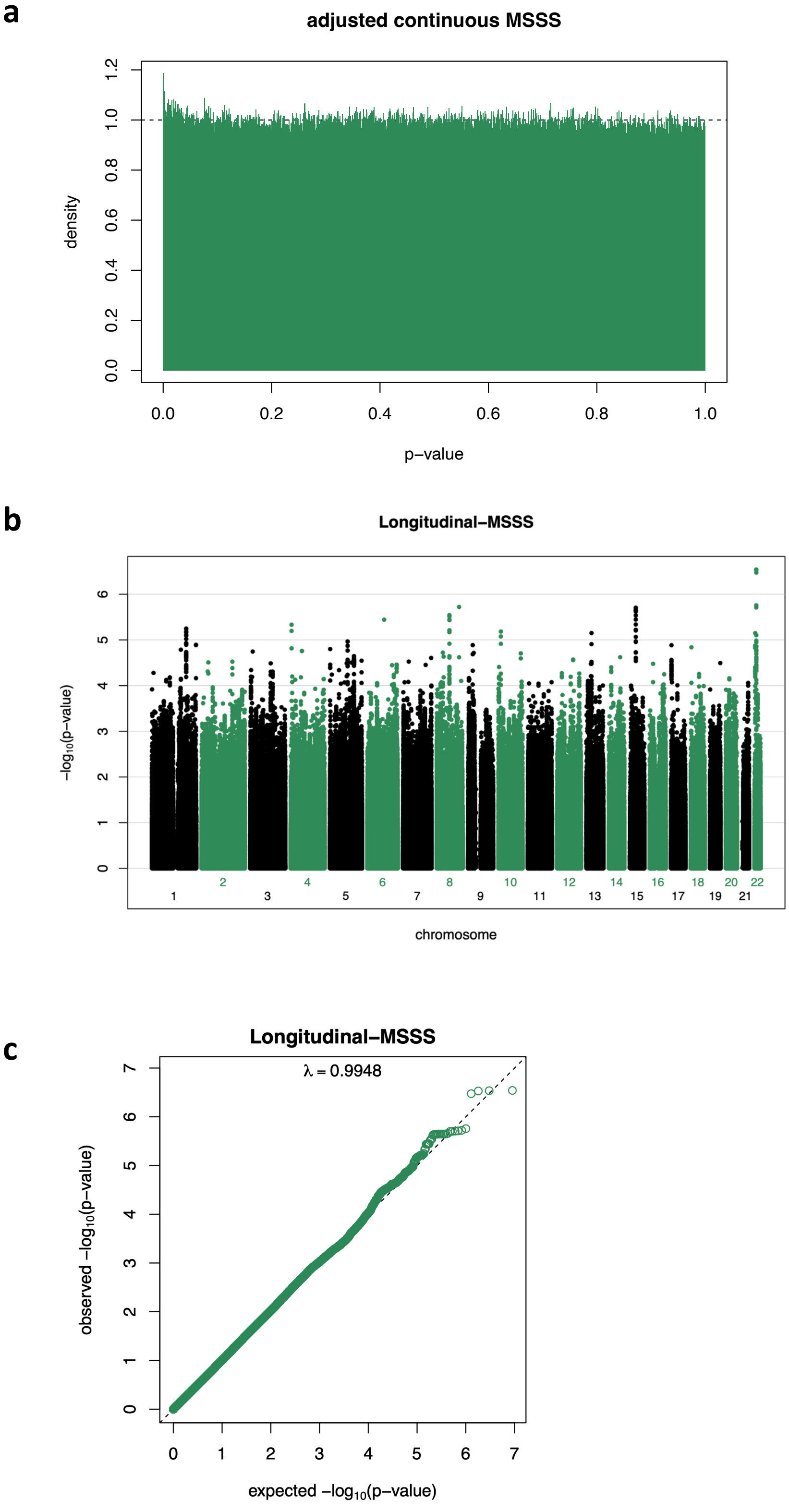

***Figure S4:*** Continuous l-MSSS GWAS adjusted fixed-effects meta-analysis **a:** p-value distribution **b:** Manhattan plot **c:** QQ plot and genomic inflation factor λ

**a**

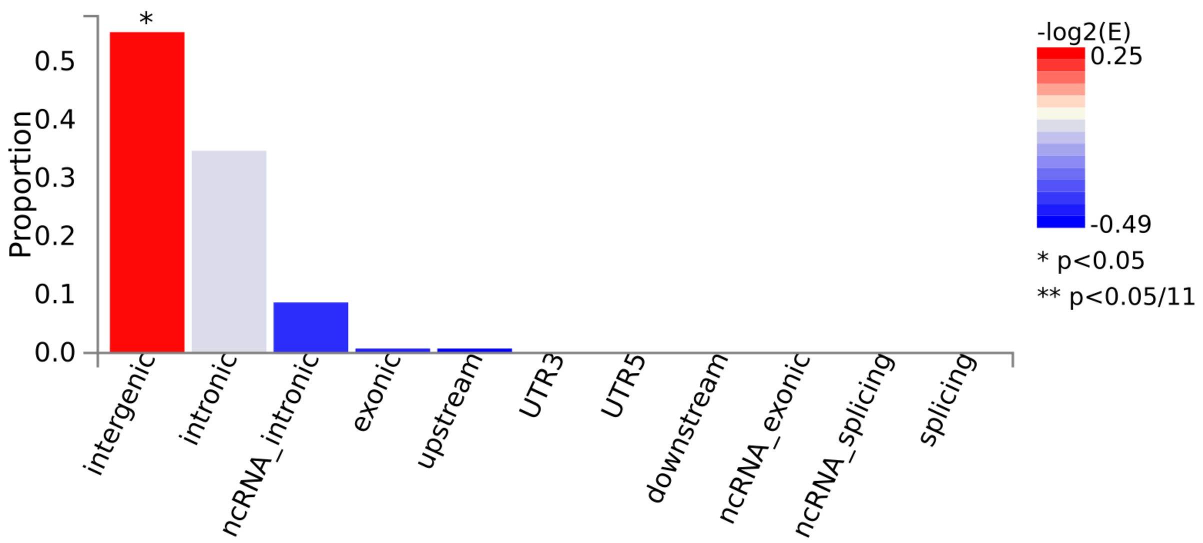

**b**

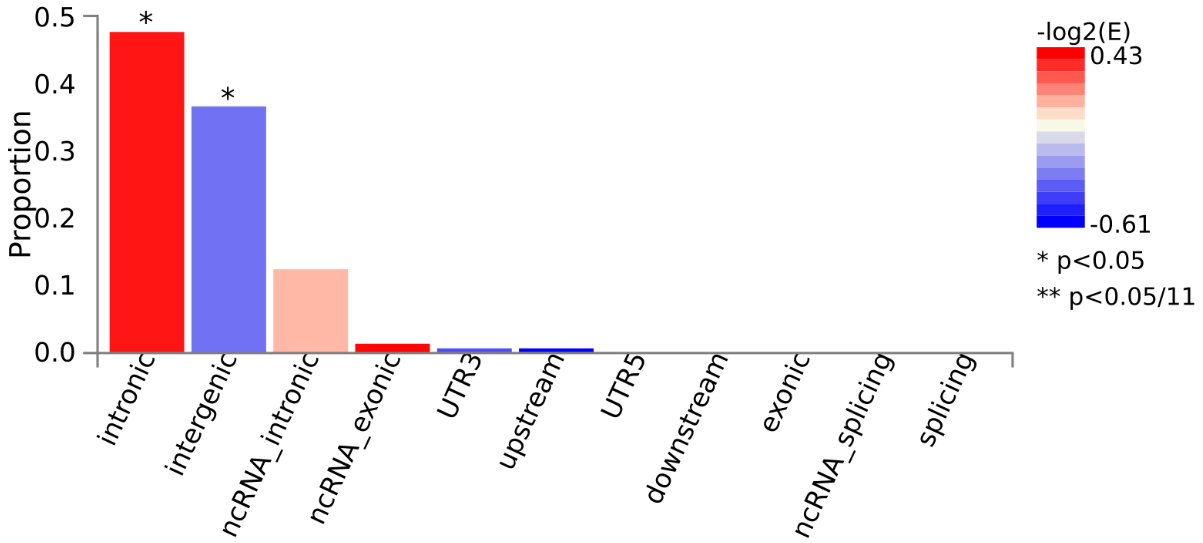

***Figure S5:*** Genomic distribution of SNVs with $p<1x10^{-5}$ in the **a:** continuous l-ARMSS phenotype GWAS analysis and **b:** continuous l-MSSS phenotype GWAS analysis as determined using FUMA.

**a**

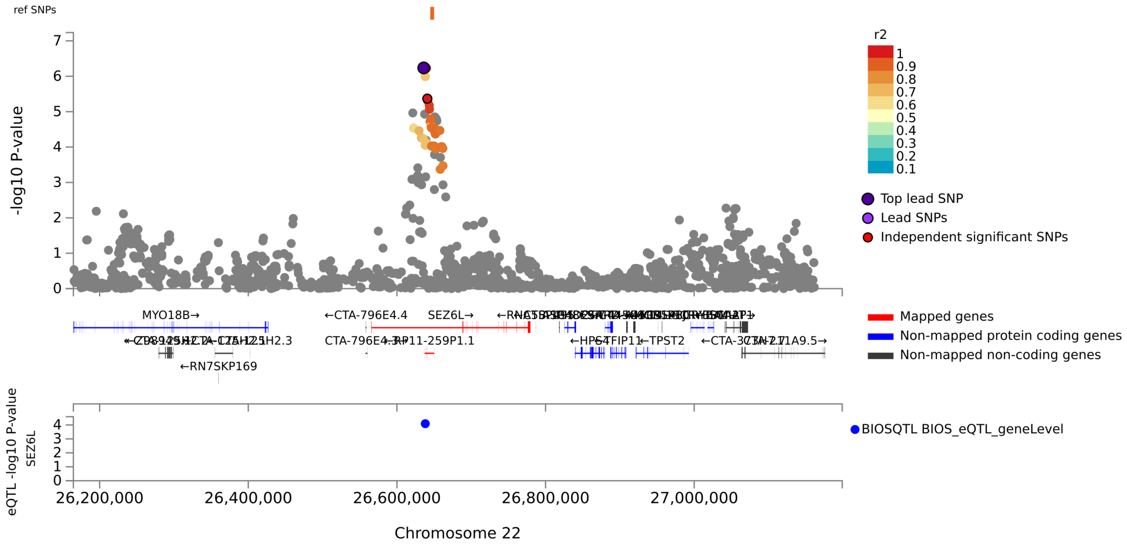

**b**

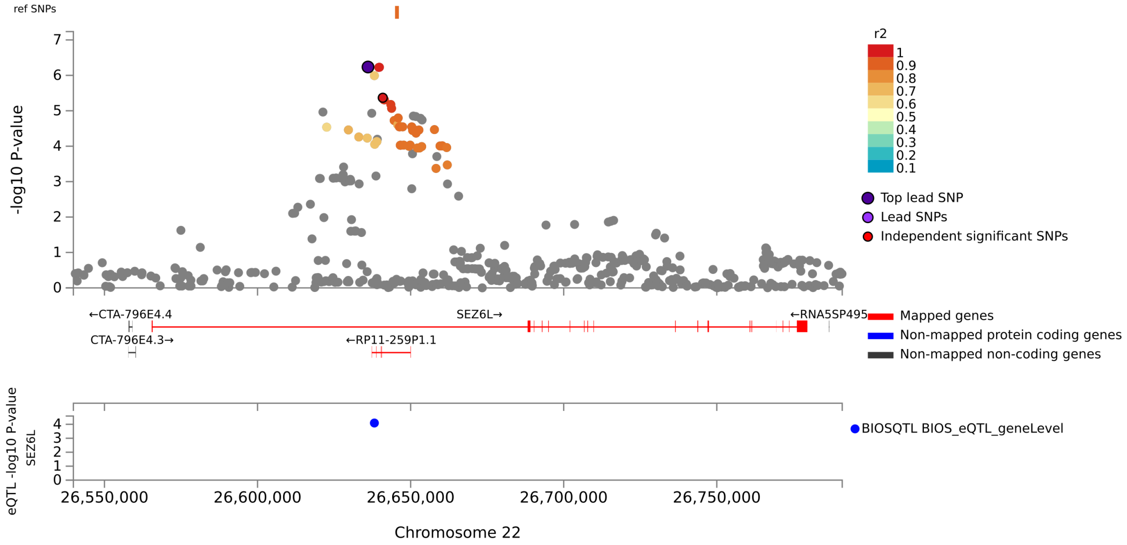

***Figure S6:*** Locus zoom plots of **a:** Chromosome 22, the region most strongly associated with longitudinal MS severity by both l-ARMSS and l-MSSS metrics. **b:** the SEZ6L gene region on Chromosome 22 demonstrates a series of SNVs in high LD with the Lead SNV (rs7289466), although independent SNVs (grey) are also present.

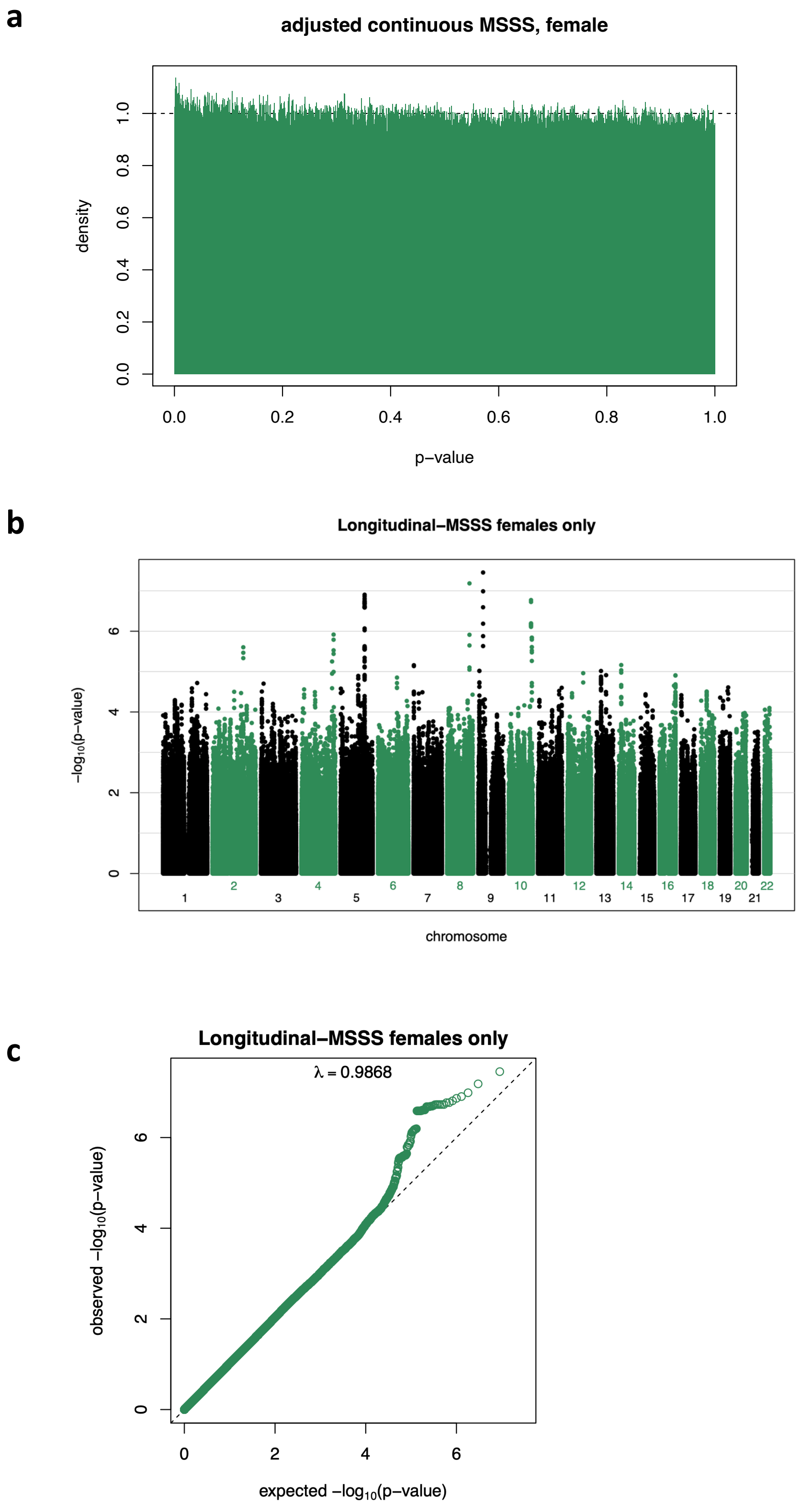

***Figure S7:*** Continuous l-MSSS GWAS in females adjusted fixed-effects meta-analysis **a:** p-value distribution **b:** Manhattan plot c: QQ plot and genomic inflation factor λ

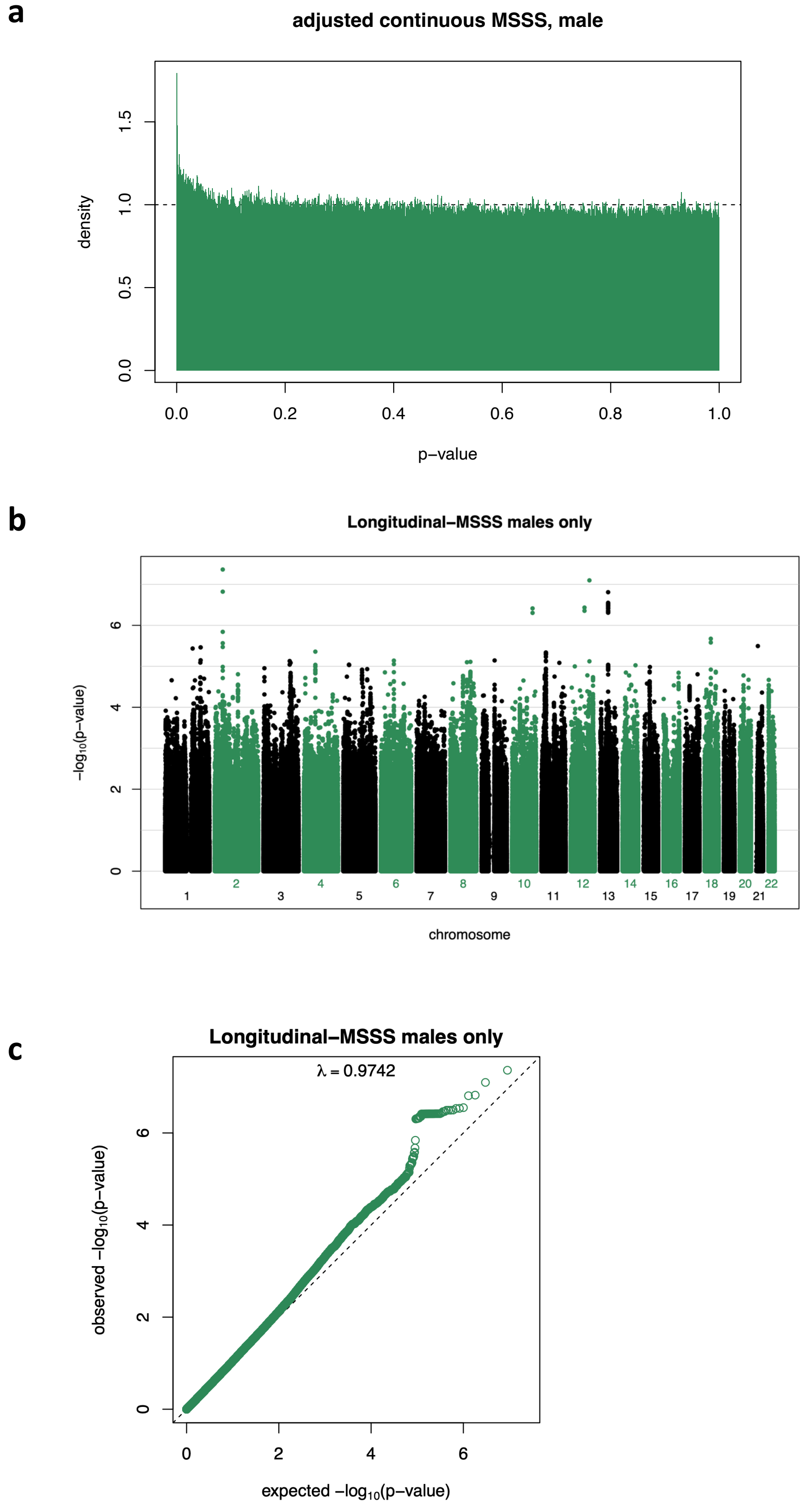

***Figure S8:*** Continuous l-MSSS GWAS in males adjusted fixed-effects meta-analysis **a:** p-value distribution **b:** Manhattan plot c: QQ plot and genomic inflation factor λ

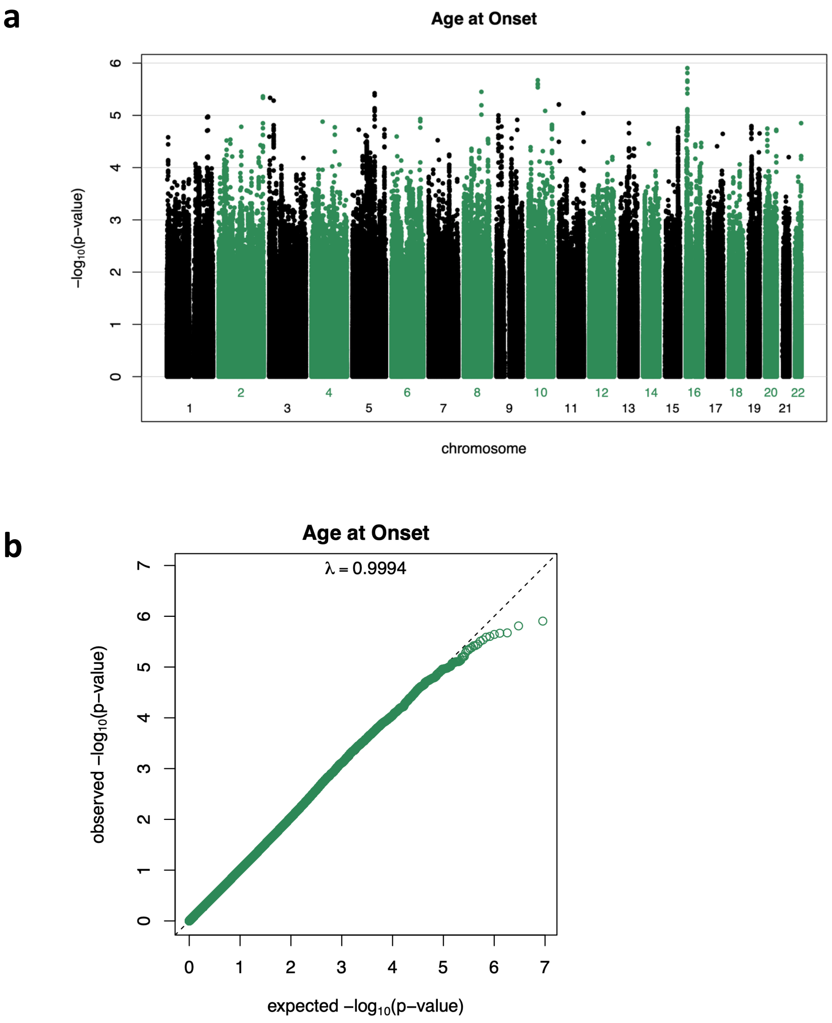

***Figure S9:*** Age at Onset adjusted fixed-effects meta-analysis **a:** Manhattan plot **b:** QQ plot and genomic inflation factor λ

**a**

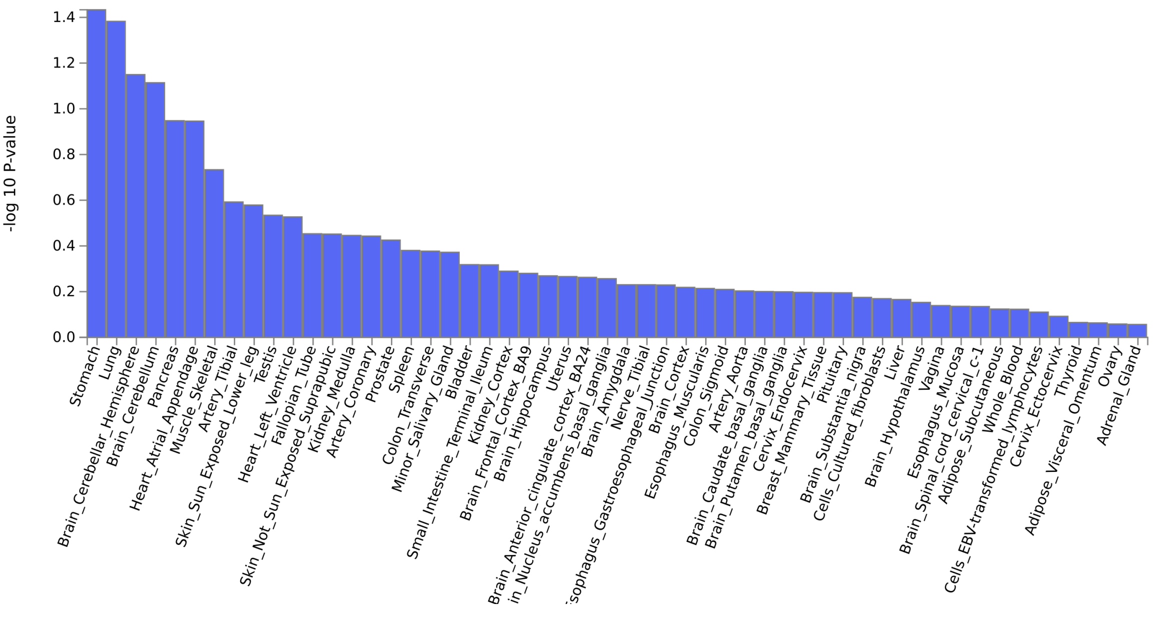

**b**

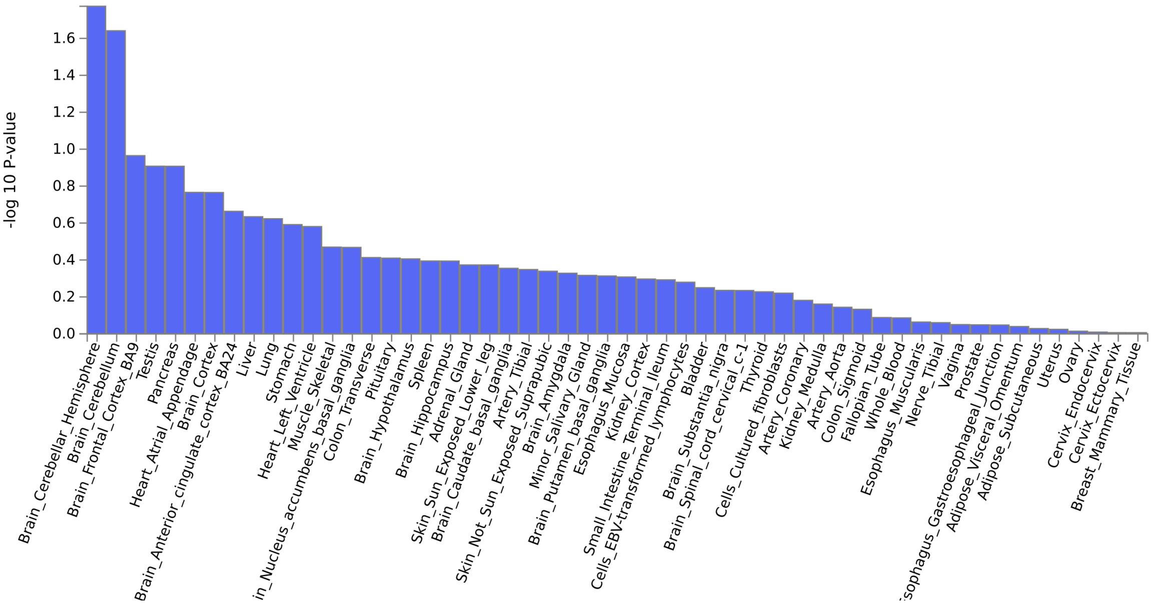

***Figure S10:*** MAGMA tissue specificity of phenotype implemented in FUMA demonstrates an overrepresentation of cerebellar hemisphere expressed genes in both a: l-ARMSS (p=0.071) and b: l-MSSS (p=0.017) GWAS analyses.

**Irreversible EDSS 3** **Irreversible EDSS 6**

**Males Only**

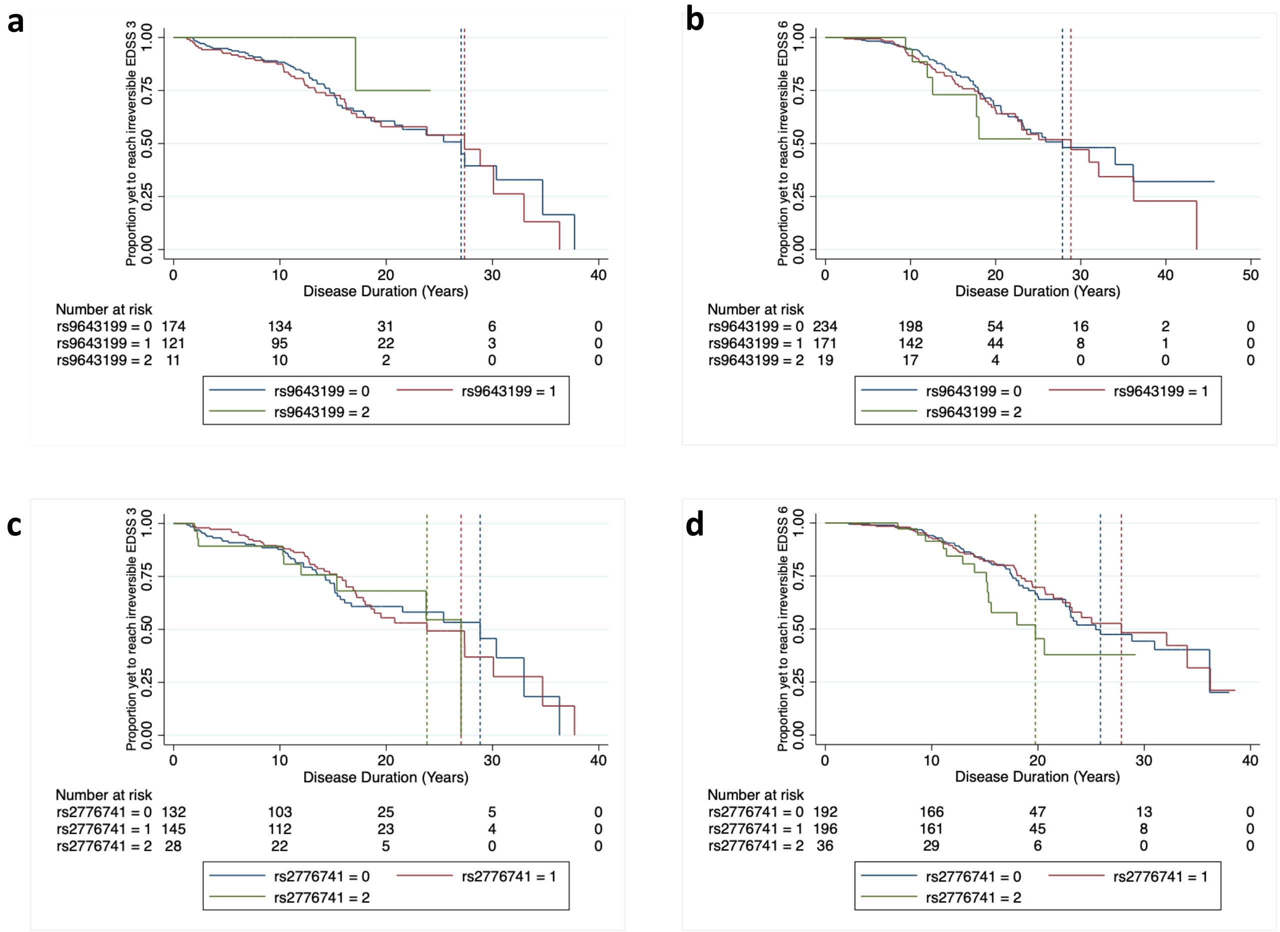

**Females Only**

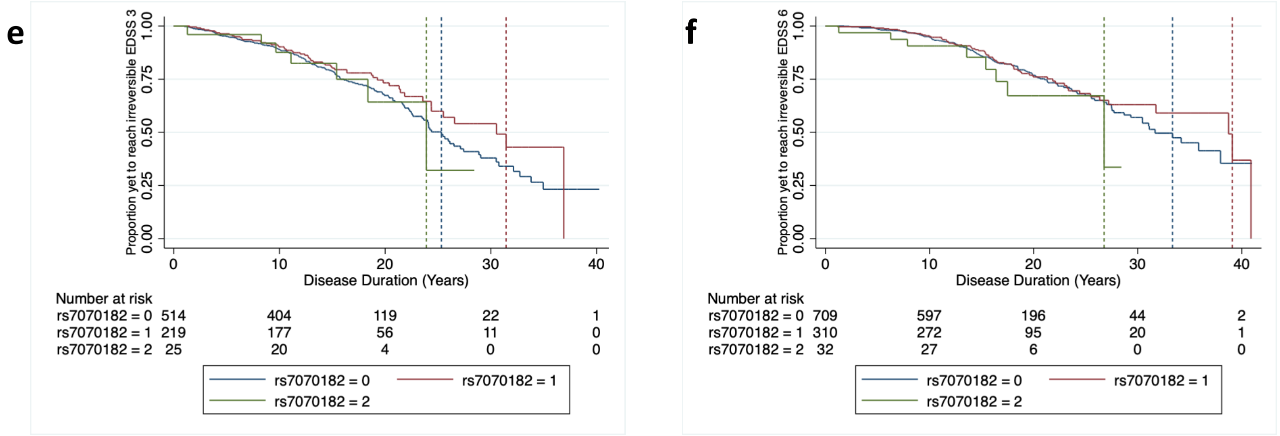

***Figure S11:*** Kaplan-Meier survival curves showing time to irreversible EDSS milestones based on the presence (1,2) or absence (0) of the minor allele at each locus **a:** rs9643199 intronic to MTSS1 time to irreversible EDSS 3 in males **b:** rs9643199 intronic to MTSS1 time to irreversible EDSS 6 in males **c:** rs2776741 near RCAN3AS time to irreversible EDSS 3 in males **d:** rs2776741 near to RCAN3AS time to irreversible EDSS 6 in males **e:** rs7070182 intronic to TCF7L2 time to irreversible EDSS 3 in females **f:** rs7070182 intronic to TCF7L2 time to irreversible EDSS 6 in females.

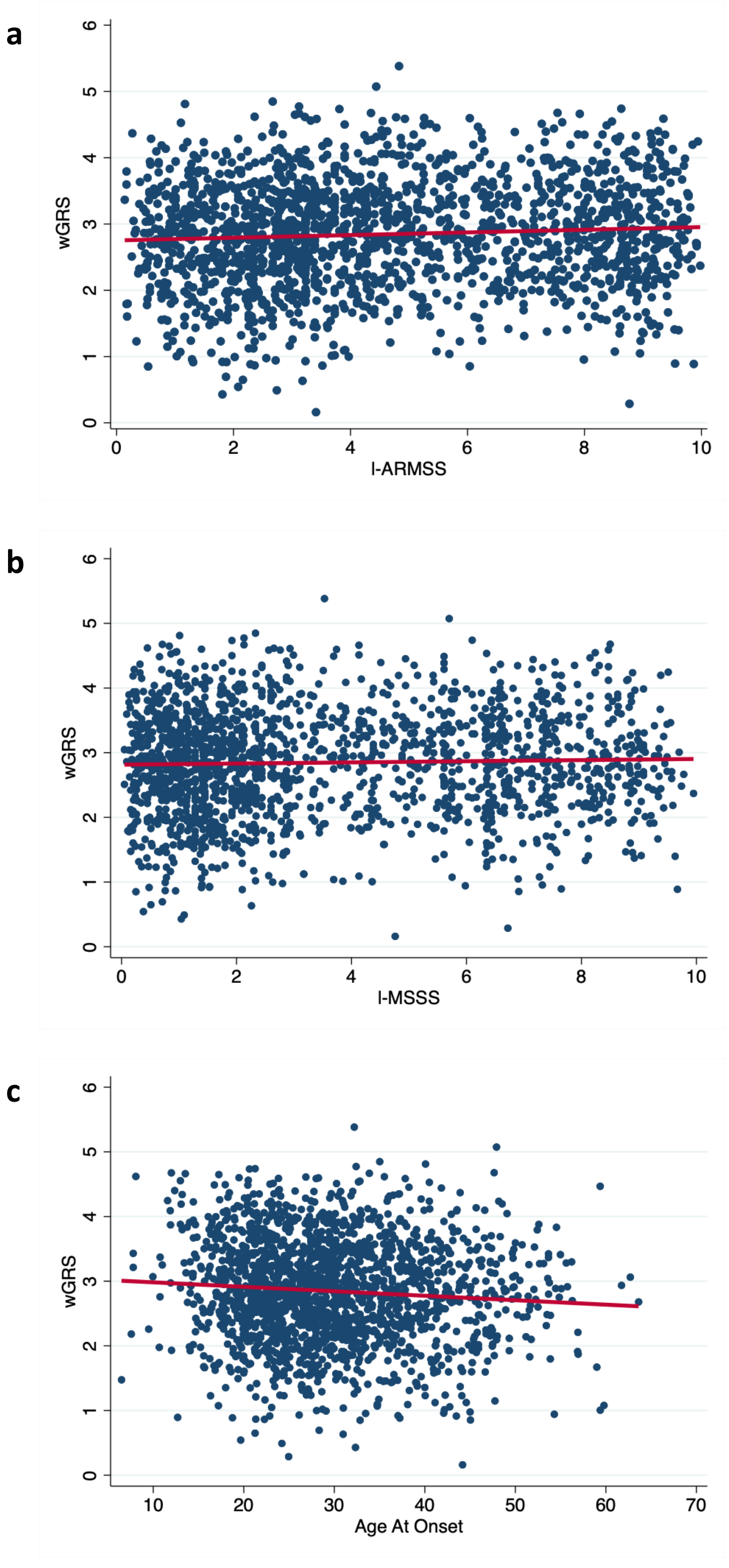

***Figure S12:*** MS susceptibility weighted genetic risk score (wGRS) plotted against **a:** l-AMRSS (r=0.07, p=0.003) **b:** l-MSSS (r=0.03, p=0.19) **c:** Age at Onset (r=-0.08, p=0.0005) demonstrates weak correlations between MS genetic risk score and phenotypes of interest.
